# Ad26.COV2.S priming provides a solid immunological base for mRNA-based COVID-19 booster vaccination

**DOI:** 10.1101/2022.07.15.22277639

**Authors:** Daryl Geers, Roos S.G. Sablerolles, Debbie van Baarle, Neeltje A. Kootstra, Wim J.R. Rietdijk, Katharina S. Schmitz, Susanne Bogers, Lennert Gommers, Nella J. Nieuwkoop, Laura L.A. van Dijk, Eva van Haren, Melvin Lafeber, Virgil A.S.H. Dalm, Abraham Goorhuis, Douwe F. Postma, Leo G. Visser, Anke L.W. Huckriede, Alessandro Sette, Alba Grifoni, Rik L. de Swart, Marion P.G. Koopmans, P. Hugo M. van der Kuy, Corine H. GeurtsvanKessel, Rory D. de Vries, the SWITCH research group

## Abstract

A large proportion of the global population received a single dose of the Ad26.COV2.S coronavirus disease-2019 (COVID-19) vaccine as priming vaccination, which was shown to provide protection against moderate to severe COVID-19. However, the emergence of severe acute respiratory syndrome coronavirus-2 (SARS-CoV-2) variants that harbor immune-evasive mutations in the spike protein led to the recommendation of booster vaccinations after Ad26.COV2.S priming. Recent studies showed that heterologous booster vaccination with an mRNA-based vaccine following Ad26.COV2.S priming leads to high antibody levels. However, how heterologous booster vaccination affects other functional aspects of the immune response remains unknown. Here, we performed immunological profiling on samples obtained from Ad26.COV2.S-vaccinated individuals before and after a homologous (Ad26.COV2.S) or heterologous (mRNA-1273 or BNT162b2) booster vaccination. Both homologous and heterologous booster vaccination increased antibodies with multiple functionalities towards ancestral SARS-CoV-2, the Delta and Omicron BA.1 variants. Especially, mRNA-based booster vaccination induced high levels of neutralizing antibodies and antibodies with various Fc-mediated effector functions such as antibody-dependent cellular cytotoxicity and phagocytosis. In contrast, T cell responses were similar in magnitude following homologous or heterologous booster vaccination, and retained functionality towards Delta and Omicron BA.1. However, only heterologous booster vaccination with an mRNA-based vaccine led to the expansion of SARS-CoV-2-specific T cell clones, without an increase in the breadth of the T cell repertoire as assessed by T cell receptor sequencing. In conclusion, we show that Ad26.COV2.S priming vaccination provides a solid immunological base for heterologous boosting with an mRNA-based COVID-19 vaccine, increasing humoral and cellular responses targeting newly emerging variants of concern.

**One sentence summary:** Ad26.COV2.S priming provides a solid immunological base for extension of cellular and humoral immune responses following an mRNA-based booster.

## Introduction

The emergence of SARS-CoV-2 variants that are antigenically distinct and can evade vaccine-induced antibody responses *(1, 2)* resulted in the recommendation of COVID-19 booster vaccinations *(3, 4)*. Currently circulating variants are predominantly viruses from the B.1.1.529 lineage (Omicron), named the BA.1-5 variants. These variants harbor several mutations in the spike (S) protein that allow for partial immune escape at the antibody level. Previous studies have shown that mRNA-based booster vaccinations increase both S-specific antibodies and to a lesser extent T cell responses, and restore clinical protection against severe disease after infection with antigenically distinct variants *(5-8)*.

According to the final evaluation of a phase 3 clinical trial, vaccination with a single dose of Ad26.COV2.S induces protection against moderate to severe-critical COVID-19, however, to varying degrees between different SARS-CoV-2 variants and the ancestral virus *(2)*. It was shown that vaccination-induced antibodies specifically have reduced reactivity to SARS-CoV-2 Omicron sub-lineages. In contrast, CD4 and CD8 T cell responses cross-react with the ancestral SARS-CoV-2 and its variants *(9)*. Compared to the mRNA-based vaccines, Ad26.COV2.S vaccination yielded lower levels of S-specific antibodies; however, these antibody levels remained stable for at least 6 months, whereas mRNA-induced antibodies waned significantly after completion of the primary regimen *(8, 10)*. Since S-specific neutralizing antibodies were identified as a correlate of protection against COVID-19 *(8, 11, 12)*, booster vaccination of Ad26.COV2.S-primed individuals was recommended to increase protection against newly emerging variants. It was previously shown that boosting Ad26.COV-2.S-primed individuals with Ad26.COV2.S, BNT162b2, or mRNA-1273 was safe and effective *(10, 13, 14)*, and that SARS-CoV-2-specific antibody and T cell responses were highest after heterologous boosting with an mRNA-based vaccine *(15)*.

SARS-CoV-2 neutralization is predominantly dependent on targeting the receptor binding domain (RBD) or N-terminal domain (NTD) of the S protein, making immune escape from neutralization possible with relatively few mutations *(16)*. Because of this, cross-neutralization of the recently emerged Omicron variants (BA.1-BA.5) is reduced or even absent in individuals who completed their primary regimen with any COVID-19 vaccine *(8, 17-24)*. However, in addition to neutralization, S-specific (non-neutralizing) antibodies can have additional effector functions by activating cellular receptors through their constant (Fc) portion. These Fc-mediated antibody functions, like antibody-dependent cellular cytotoxicity (ADCC) and antibody-dependent cellular phagocytosis (ADCP), have been associated with reduced COVID-19 severity and mortality *(25, 26)*. Notably, ADCC-mediating antibodies were also identified as a correlate of protection against other respiratory viral infections such as respiratory syncytial virus (RSV), influenza virus and human immunodeficiency virus (HIV) infections *(27-29)*. Since non-neutralizing antibodies can potentially bind epitopes spanning the entire SARS-CoV-2 S protein, including more conserved regions in the S2 domain, they mediate broader cross-reactivity to emerging SARS-CoV-2 variants *(30-34)*. Considering the high number of mutations in the S protein of the Omicron sub-lineage, it is important to assess the capacity of antibodies that induce non-neutralizing functions to cross-recognize SARS-CoV-2 variants.

In addition to SARS-CoV-2-specific antibody responses, virus-specific CD4 and CD8 T cells play an important role in controlling a SARS-CoV-2 infection *(35-37)*, mainly by clearing virus-infected cells and thereby limiting disease severity *(11)*. All vaccines approved in Europe, including Ad26.COV2.S, were shown to induce virus-specific CD4 and CD8 T cells *(6, 8, 38, 39)*, which remained stable in magnitude and functionality over time and retained cross-reactivity with variants, including the Omicron BA.1 variant *(8, 9, 30, 40, 41)*. However, how booster vaccinations in Ad26.COV2.S primed individuals affect the magnitude, breadth, and diversity of the T cell response remains elusive. Booster vaccination may induce novel T cell clones targeting conserved epitopes between emerging variants and the ancestral SARS-CoV-2.

Here, we performed immunological profiling of SARS-CoV-2-specific antibody and T-cell responses to ancestral SARS-CoV-2, Delta and Omicron BA.1 variants in health care workers (HCW) primed with Ad26.COV2.S and boosted with a homologous or heterologous mRNA-based vaccine. To assess the immunological foundation laid by Ad26.COV2.S priming, immune responses were assessed pre-booster vaccination (3 months after priming), and 28 days after homologous or heterologous booster vaccination.

## Results

### Cohort description

For the characterization of SARS-CoV-2-specific immune responses before and 28 days after homologous or heterologous booster vaccination in Ad26.COV2.S primed individuals, n = 60 study participants were randomly selected based on availability of samples from n = 434 healthcare workers (HCW) from the previously reported SWITCH trial *(10)*. Of the 60 HCW included, n = 15 received a second vaccination with Ad26.COV2.S, n = 15 received mRNA-1273, n = 15 received BNT162b2, and n= 15 did not receive a second vaccination (no boost). Participants received their second vaccination ±95 days after priming with Ad26.COV2.S. The study design is shown in **Suppl. Fig. 1A** and participant characteristics are summarized in **Table 1**. At baseline, before booster vaccination, there was no difference in binding antibody levels **(Suppl. Fig. 1B)** and T cell responses measured in whole blood **(Suppl. Fig. 1C)** across groups. Groups did not differ in female to male composition from our original study; however, there was a significant age difference. Participants from the heterologous vaccination regimens had a mean age of 36 or 37 years for BNT162b2 and mRNA-1273 vaccination, respectively. In contrast, Ad26.COV2.S boosted participants had a mean age of 51 years.

**Table 1:** baseline characteristics.

|  |  | Total<br>N= 60 | Ad26.COVS2.S/ no boost<br>N= 15 | Ad26.COVS2.S/<br>Ad26.COVS2.S<br>N= 15 | Ad26.COVS2.S/ mRNA-1273<br>N= 15 | Ad26.COVS2.S/ BNT162b2<br>N= 15 | *p value* |
| --- | --- | --- | --- | --- | --- | --- | --- |
| **Sex** | Male | 20 (33) | 5 (33) | 6 (40) | 2 (13) | 7 (47) | *0.25* |
|  | Female | 40 (67) | 10 (67) | 9 (60) | 13 (87) | 8 (53) |  |
| **Age** |  | 41.5 [31.8-51.0] | 51.0 [38.5-55.5] | 51.0 [38.5-55.0] | 37.0 [28.5-43.5] | 36.0 [31.0-40.0] | ***0.007*** |
| **BMI** |  | 24.1 [21.1-26.6] | 24.2 [21.7-26.6] | 23.3 [20.8-24.6] | 21.5 [20.6-25.4] | 25.9 [22.0-27.3] | *0.35* |
| **Time between first vaccination and SV1** |  | 95.5 [87.5-98.8] | 98.4 [88.0-99.5] | 95.6 [89.5-97.9] | 90.4 [87.0-98.0] | 94.5 [81.0-98.5] | *0.54* |
| **Time between SV1 and SV2** |  | 27.5 [27.4-27.6] | 27.6 [27.5-27.6] | 27.5 [27.5-27.6] | 27.5 [27.5-27.8] | 27.5 [27.5-28.1] | *0.93* |
| **Immunogenicity data** |  |  |  |  |  |  |  |
| **Liaison - SARS-CoV-2 spike protein (S)-specific binding antibodies (BAU/ml)** | **SV1 - before boost** | 111.0 [54.1-212.0] | 178.0 [50.6 - 375.5] | 103.0 [71.1-194.0] | 91.4 [61.5-140.5] | 147.0 [54.9-309.5] | *0.82* |
|  | **SV2 - 28 days after boost** | 1270.0 [289.8-2962.5] | 200.0 [44.5-333.0] | 466.0 [280.5-720.0] | 5050.0 [2545.0 - 7360.0] | 2680.0 [1640.0-3965.0] | ***<0.001*** |
| **IGRA - S-specific T-cell response (IU/ml)** | **SV1 - before boost** | 0.24 [0.07-0.75] | 0.16 [0.05-0.52] | 0.18 [0.07-0.35] | 0.49 [0.05-1.36] | 0.38 [0.19-0.98] | *0.29* |
|  | **SV2 - 28 days after boost** | 0.64 [0.13-1.58] | 0.09 [0.02-0.29] | 0.26 [0.13-0.56] | 1.43 [0.87-2.73] | 1.03 [0.85-2.42] | ***<0.001*** |
| Note: Values are number (percentage) for categorical variables and median [interquartile range] for continuous variables. SV1= study visit 1 (prior to second vaccination); SV2 = study visit 2 (potential second vaccination) |  |  |  |  |  |  |  |
| Statistics: Fisher exact test was used to assess differences between categorical variables and Kruskal Wallis test between continuous variables |  |  |  |  |  |  |  |

### Binding antibodies cross-react with the Delta and Omicron variants

Binding antibodies to ancestral SARS-CoV-2, and the Delta, or Omicron BA.1 variant S proteins were assessed by ELISA **(Fig. 1A)**. A significant increase in binding antibody levels was observed 28 days after both homologous and heterologous booster vaccination **(Fig. 1B and Suppl. Fig. 2)**. We found the lowest binding antibody titer in the no boost group (GMT of 1192). The binding antibody titers were higher after homologous (Ad26.COV2.S; GMT of 3774) and particularly after heterologous booster with mRNA-1273 (GMT of 117660) or BNT162b2 (GMT of 58747) **(Fig. 1B)**. These patterns were comparable with previously reported S1-specific binding antibodies as measured by Liaison **(Suppl. Fig. 1B)** *(10)*. We found that binding antibodies were in general cross-reactive with both the Delta and Omicron BA.1 variant S proteins, although significantly lower antibody titers were found against the Omicron S protein across all groups and timepoints **(Fig. 1C)**. Although, lower levels of binding antibodies to the Delta variant S-protein were also observed following Ad26.COV2.S and BNT162b2 booster vaccination, overall no significant differences were observed. To further analyze these differences in antibody production/level at cellular level, we determined the percentage of total RBD-specific B cells by flow cytometry in peripheral blood mononuclear cells (PBMC). Ancestral RBD-specific B cells were detected in the pre-booster samples of all participants and no differences were observed between the groups. Interestingly, booster vaccination with either Ad26.COV2.S, mRNA-1273 or BNT162 did not increase the frequency, nor change the phenotype of RBD-specific B cells. Similar frequencies of RBD-specific B cells, RBD-specific memory B cells as well as RBD-specific IgG memory B cells were observed pre- and post-booster with all vaccination regimens **(Suppl. Fig. 3)**.

**Figure 1:**
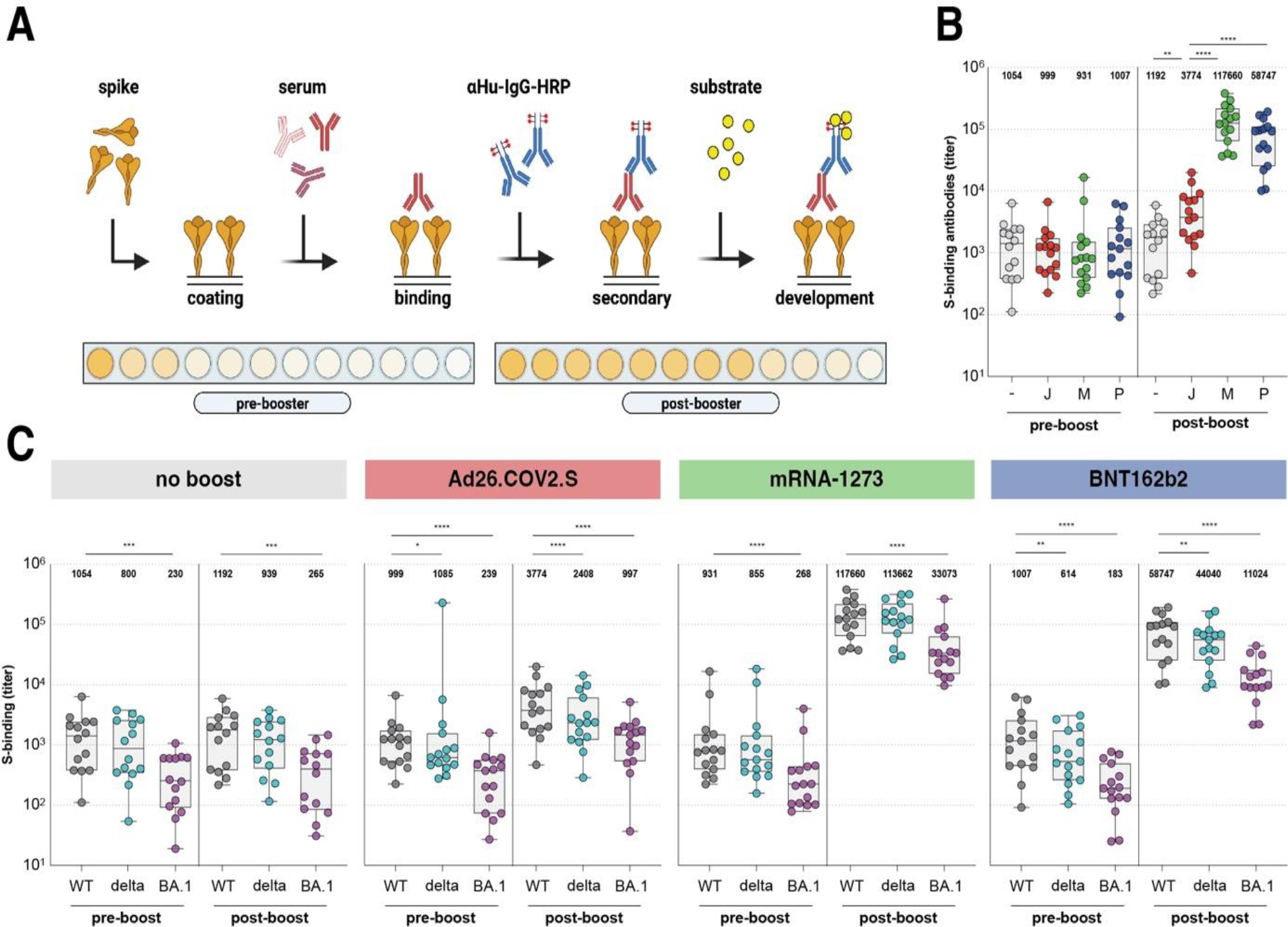
Binding antibodies are boosted by homologous or heterologous vaccination, but bind less to the Omicron BA.1 variant. **(A)** Enzyme-linked immunosorbent assay (ELISA) methodology. **(B)** Binding antibodies pre- and post-booster vaccination after no boost (grey), Ad26.COV2.S boost (red), mRNA-1273 boost (green), or BNT162b2 boost (blue). Geometric mean titers (GMT) are depicted above the graph. **(C)** Binding antibodies against ancestral SARS-CoV-2 (grey), Delta (cyan), or Omicron BA.1 (pink) variants pre- and post-booster. GMT are depicted for each group. S = spike protein, - = no boost, J = Ad26.COV2.S, M = mRNA-1273, P = BNT162b2, WT = ancestral virus, delta = Delta variant, BA.1 = Omicron BA.1 variant. Symbols represent individual donors. Box plot depicts the median with range (min to max). p <0.05 (*), p <0.01 (**), p <0.001 (***), and p <0.0001 (****).

### Antibodies with Fc-mediated functions cross-react with the Delta and Omicron variants

Two different antibody effector functions mediated by the Fc region were assessed: the induction of ADCC and ADCP. ADCC-mediating antibodies were measured in a functional NK cell degranulation assay performed on S protein coated plates **(Fig. 2A)**. Similar to the binding antibodies, higher levels of ADCC-mediating antibodies were observed after Ad26.COV2.S booster vaccination (median of 16%) compared to no boost (median of 9.5%). The highest levels of ADCC-mediating antibodies were observed after mRNA-1273 (median 20.0%) or BNT162b2 (median of 20%) booster vaccination **(Fig. 2B)**. Although ADCC-mediating antibodies cross-reactive with the Delta variant S protein were detected in all groups at all timepoints, these were significantly lower compared to antibodies against the ancestral S protein **(Fig. 2C)**. In contrast to what was observed with binding antibodies, ADCC-mediating antibodies cross-reactive with the Omicron S protein were hardly detected, and could only be measured after mRNA-1273 (median of 11%) or BNT162b2 (median of 11%) booster vaccination **(Fig. 2C)**. Additionally, we measured ADCP-mediating antibodies in a functional THP-1 phagocytosis assay with ancestral S protein coated beads **(Fig. 2D)**. Similarly to ADCC-mediating antibodies, Fc-mediated phagocytosis was boosted by both homologous or heterologous vaccination, and highest after mRNA-1273 (GMT of 41438) or BNT162b2 (GMT of 45788) booster vaccination as compared to Ad26.COV2.S (GMT of 3373) vaccination **(Fig. 2E)**. Representative dilution series and individual dilution series per vaccination regimen are shown in **Suppl. Fig. 4A and 4B**, respectively.

**Figure 2:**
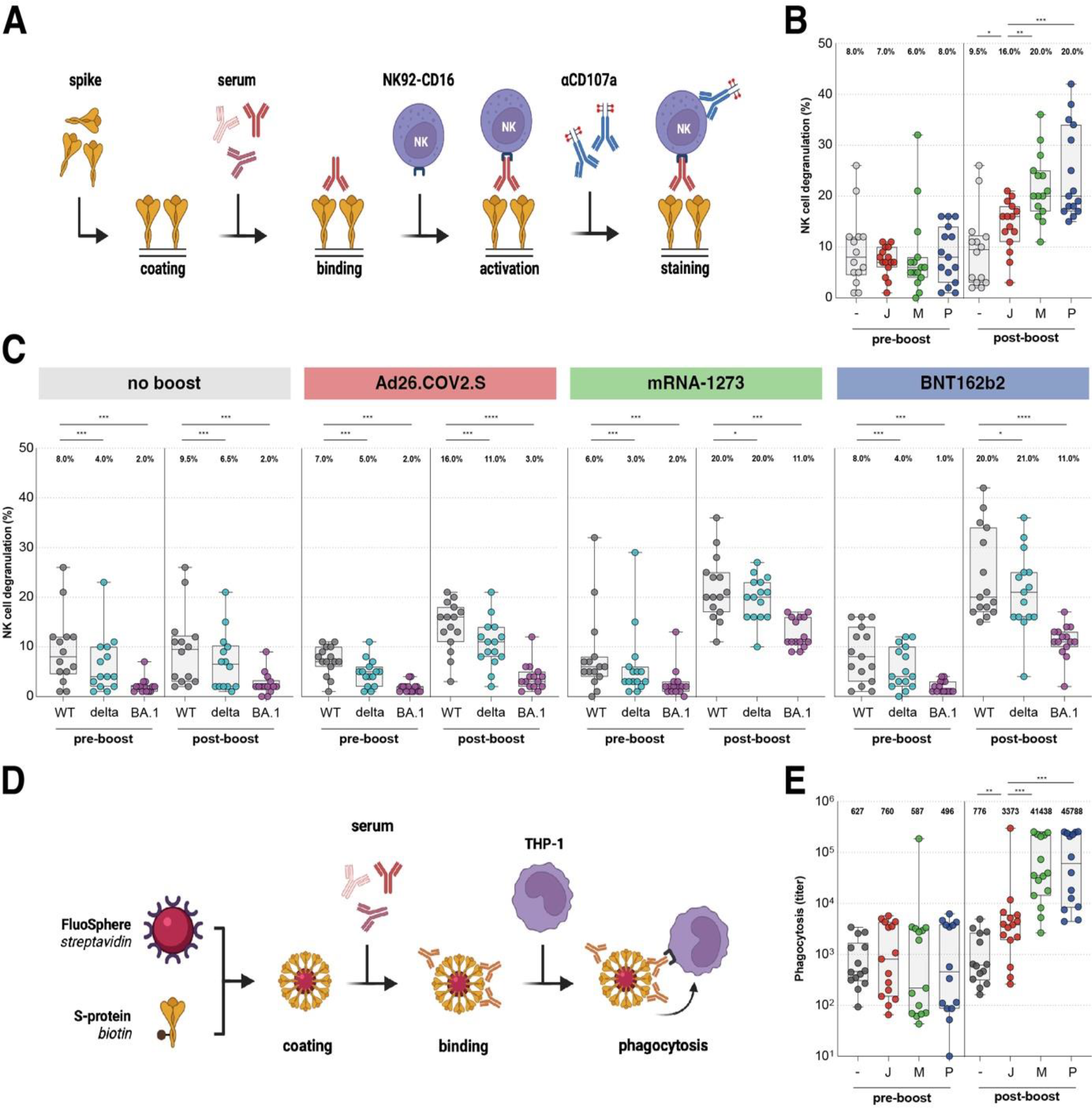
Fc-mediated antibody functions are boosted by homologous or heterologous vaccination, but less functional against the Omicron BA.1 variant. **(A)** Antibody-dependent cell-mediated cytotoxicity (ADCC) assay methodology. **(B)** NK cell degranulation (%) to the ancestral SARS-CoV-2 pre- and post-booster vaccination after no boost (grey), Ad26.COV2.S boost (red), mRNA-1273 boost (green), or BNT162b2 boost (blue). Median percentages are depicted above graph. **(C)** NK cell degranulation to ancestral SARS-CoV-2 (grey), Delta (cyan), or Omicron BA.1 (pink) variants pre- and post-booster vaccination. Median percentages are depicted above graph. **(D)** Antibody-dependent cell-mediated phagocytosis (ADCP) assay methodology. **(E)** Phagocytosis-mediating antibodies to ancestral SARS-CoV-2 pre- and post-booster vaccination. Geometric mean titers (GMT) are depicted above graph. - = no boost, J = Ad26.COV2.S, M = mRNA-1273, P = BNT162b2, WT = ancestral virus, delta = Delta variant, BA.1 = Omicron BA.1 variant. Symbols represent individual donors. Box plot depicts the median with range (min to max). p <0.05 (*), p <0.01 (**), p <0.001 (***), and p <0.0001 (****).

### Cross-neutralization of the Omicron BA.1 is increased after heterologous booster

Neutralizing antibodies were assessed in an infectious virus neutralization assay with the ancestral SARS-CoV-2, Delta, and Omicron BA.1 variants **(Fig. 3A)**. As reported previously, mRNA-based booster vaccination with either mRNA-1273 or BNT162b2 led to the highest neutralizing antibodies against the ancestral SARS-CoV-2, GMT of 3983 and GMT 3382, respectively *(10)* **(Fig. 3B)**. Cross-neutralizing antibodies against Delta and Omicron BA.1 were observed after mRNA-based booster vaccination, although at a significantly lower level compared to the ancestral SARS-CoV-2. Strikingly, cross-neutralization of the Omicron BA.1 variant was virtually absent (GMT of 13) after Ad26.COV2.S booster vaccination **(Fig. 3C)**. Individual S-curves per vaccination regimen are shown in **Suppl. Fig. 5**. We examined the correlations between S-specific binding antibodies and S1-binding antibodies **(Fig. 4A)**, and their functionalities including neutralization (PRNT50) **(Fig. 4B)**, NK cell degranulation (ADCC) **(Fig. 4C)**, and phagocytosis (ADCP) **(Fig. 4D)** against the ancestral SARS-CoV-2 and found all correlations to be positive and significant (p<0.05).

**Figure 3:**
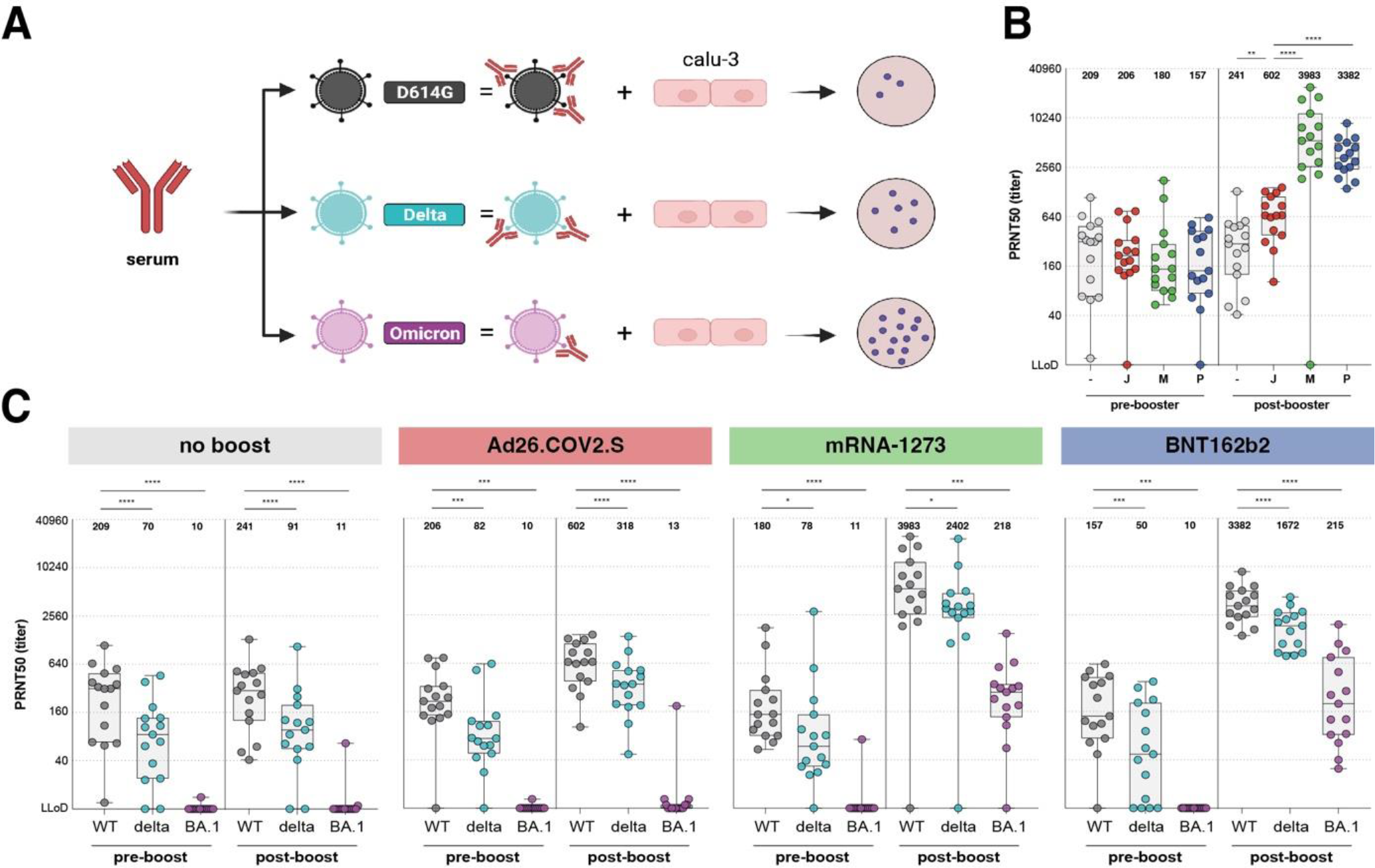
Neutralizing antibodies are boosted by homologous or heterologous vaccination, and cross-neutralize the Omicron BA.1 variant. **(A)** Plaque-reduction neutralization test (PRNT) assay methodology. **(B)** PRNT50 titer to ancestral SARS-CoV-2 pre- and post-booster vaccination after no boost (grey), Ad26.COV2.S boost (red), mRNA-1273 boost (green), or BNT162b2 boost (blue). Geometric mean titers (GMT) are depicted above graph. **(C)** PRNT50 titer pre- and post-booster vaccination for ancestral SARS-CoV-2 (grey), Delta (cyan), or Omicron BA.1 (pink) variants. Geometric mean titers (GMT) are depicted above graph. PRNT50 = plaque reduction neutralization test 50% end-point, - = no boost, J = Ad26.COV2.S, M = mRNA-1273, P = BNT162b2, WT = ancestral virus, delta = Delta variant, BA.1 = Omicron variant. Symbols represent individual donors. Box plot depicts the median with range (min to max). p <0.05 (*), p <0.01 (**), p <0.001 (***), and p <0.0001 (****).

**Figure 4:**
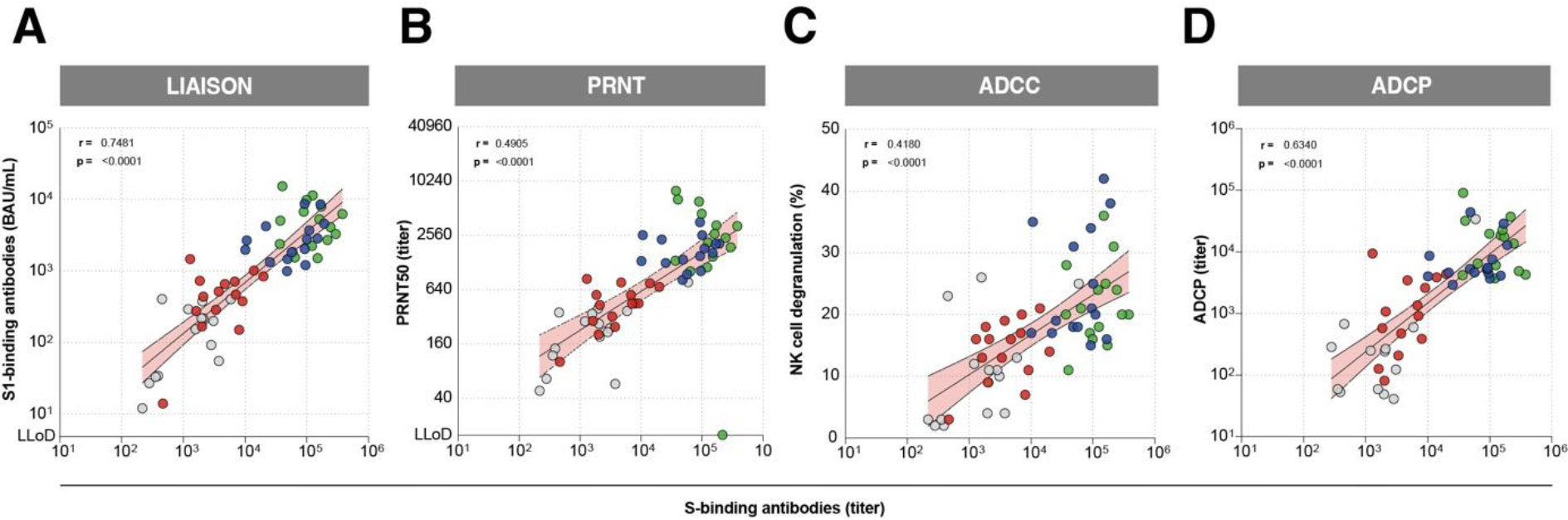
Functional antibody responses correlate with antibody binding to the ancestral SARS-CoV-2. **(A)** Correlation between S1-specific binding antibodies measured by Liaison and S-specific binding antibodies measured by ELISA. **(B)** Correlation between neutralizing antibodies and S-specific binding antibodies measured by ELISA **(C)** Correlation between NK cell degranulation mediating S-specific antibodies (ADCC) and S-specific antibody binding measured by ELISA. **(D)** Correlation between phagocytosis-mediating antibody titers (ADCP) and S-specific binding antibodies as measured by ELISA. Colors represent different booster groups: no boost (grey), Ad26.COV2.S boost (red), mRNA-1273 boost (green), and BNT162b2 boost (blue). Symbols represent individual donors post-booster vaccination. Simple linear regression analysis on log-transformed data was used to calculate Pearson’s correlation coefficient and p-values.

### S-specific CD4 and CD8 T cells cross-react with both the Delta and Omicron variant

Further, we measured T cell responses before and after homologous or heterologous booster vaccination. To directly assess T cell responses in whole blood, we previously performed an interferon gamma (IFNγ) release assay (IGRA), and found that T cell responses were boosted by both homologous and heterologous booster vaccination *(10)* **(Suppl. Fig. 1B)**. To asses T cell responses in depth, PBMCs were stimulated with overlapping peptide pools spanning the full-length ancestral S-protein and responses were measured via IFN-γ ELISPOT **(Fig. 5A)**. Here, we found that mRNA-1273 booster vaccination induced higher numbers of IFN-γ producing T cells to ancestral SARS-CoV-2, with a significantly higher T cell response following mRNA-1273 booster vaccination **(Fig. 5B)**.

**Figure 5:**
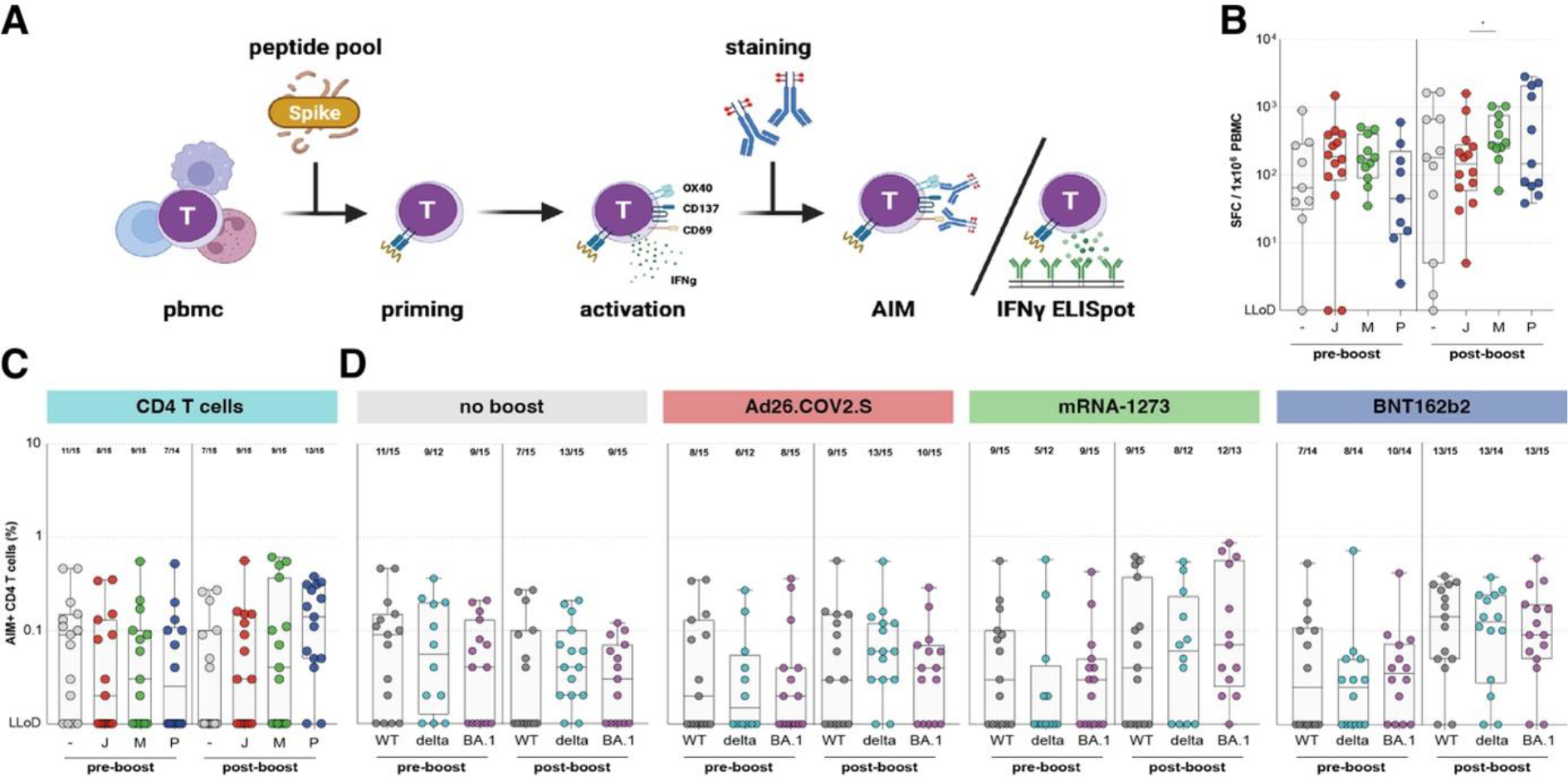
SARS-CoV-2-specific CD4 and CD8 T cells cross-react with Omicron BA.1. **(A)** Activation induced marker (AIM) assay and IFN-ɣ ELISPOT methodology. **(B)** IFN-³ secreting T cells after stimulation with an overlapping S peptide pool from the ancestral SARS-CoV-2 pre- and post-booster vaccination, after no boost (grey), Ad26.COV2.S boost (red), mRNA-1273 boost (green), or BNT162b2 boost (blue). **(C)** Comparison of CD4 T cell responses to ancestral SARS-CoV-2. **(D)** CD4 T cell responses against ancestral SARS-CoV-2 (grey), Delta (cyan), or Omicron BA.1 (pink) variants pre- and post-booster vaccination. - = no boost, J = Ad26.COV2.S, M = mRNA-1273, P = BNT162b2, WT = ancestral virus, delta = Delta variant, BA.1 = Omicron variant. Symbols represent individual donors. Box plot depicts the median with range (min to max). p <0.05 (*), p <0.01 (**), p <0.001 (***), and p <0.0001 (****).

To differentiate between CD4 and CD8 T cell responses, and to measure variant-specific responses, PBMCs were stimulated with overlapping peptide pools representing the full-length S protein from the ancestral SARS-CoV-2, and the Delta and Omicron BA.1 variants **(Fig. 5A)**. Following stimulation, CD4 (OX40^+^CD137^+^) and CD8 (CD69^+^CD137^+^) T cell activation induced marker (AIM) expression was measured by flow cytometry **(Suppl. Fig. 6A)**. CD4 and CD8 T cell responses were detected in 32/60 (53%) of participants pre-booster, and levels were comparable between groups. Booster vaccination with either Ad26.COV2.S or mRNA-1273 did not significantly increase CD4 T cell responses. Interestingly, booster vaccination with BNT162b2 increased the number of participants with a measurable CD4 T cell response from 7/14 (50%) to 13/15 (87%), with a significantly higher percentage of activated CD4 T cells (GM of 0.03% to 0.1%) after booster vaccination. In contrast, CD4 T cell responses waned for the no boost group (11/15 responders at baseline to 7/15 responders 28 days later) **(Fig. 5C)**. CD4 T cell reactivity with the Delta and Omicron BA.1 variant was maintained, and comparable to reactivity with the ancestral S protein **(Fig. 5D**). As for CD8 T cell responses, no clear boosting effect of either homologous or heterologous vaccination was observed on basis of number of responders (**Suppl. Fig. 6B)**. Similar to CD4 T cells, CD8 T cells equally reacted with all SARS-CoV-2 variants tested **(Suppl. Fig. 6C)**.

### mRNA-based booster vaccination led to expansion of S-specific T cell clones

We evaluated the expansion, breadth and depth of the SARS-CoV-2-specific T cell response after different booster regimens. TCRβ sequencing was performed to define the repertoires of N=30 participants (N=7 no boost, N=7 Ad26.COV2.S boost, N=10 mRNA-1273 boost, and N=6 BNT162b2 boost) pre- and post-booster vaccination *(42)*. Initially, we compared pre- and post-booster vaccination within donors to identify expanding clones after booster vaccination (representative example shown in **Suppl. Fig. 7A**). The number of expanding clones ranged from 0-98 across subjects. In a time period of 28 days a ‘background’ expansion of 3-5 clones is expected, similar to what is observed in participants that did not receive a boost **(Fig. 6A and Suppl. Fig. 7B)**. More expanding clones were observed in the Ad26.COV2.S-boosted individuals as compared to no boost (dominated by 73 expanding clones in 1 individual), but especially in the mRNA-1273 and BNT162b2-boosted individuals the number of expanding clones was in general >20 **(Fig. 6A and Suppl. Fig. 7B)**.

**Figure 6:**
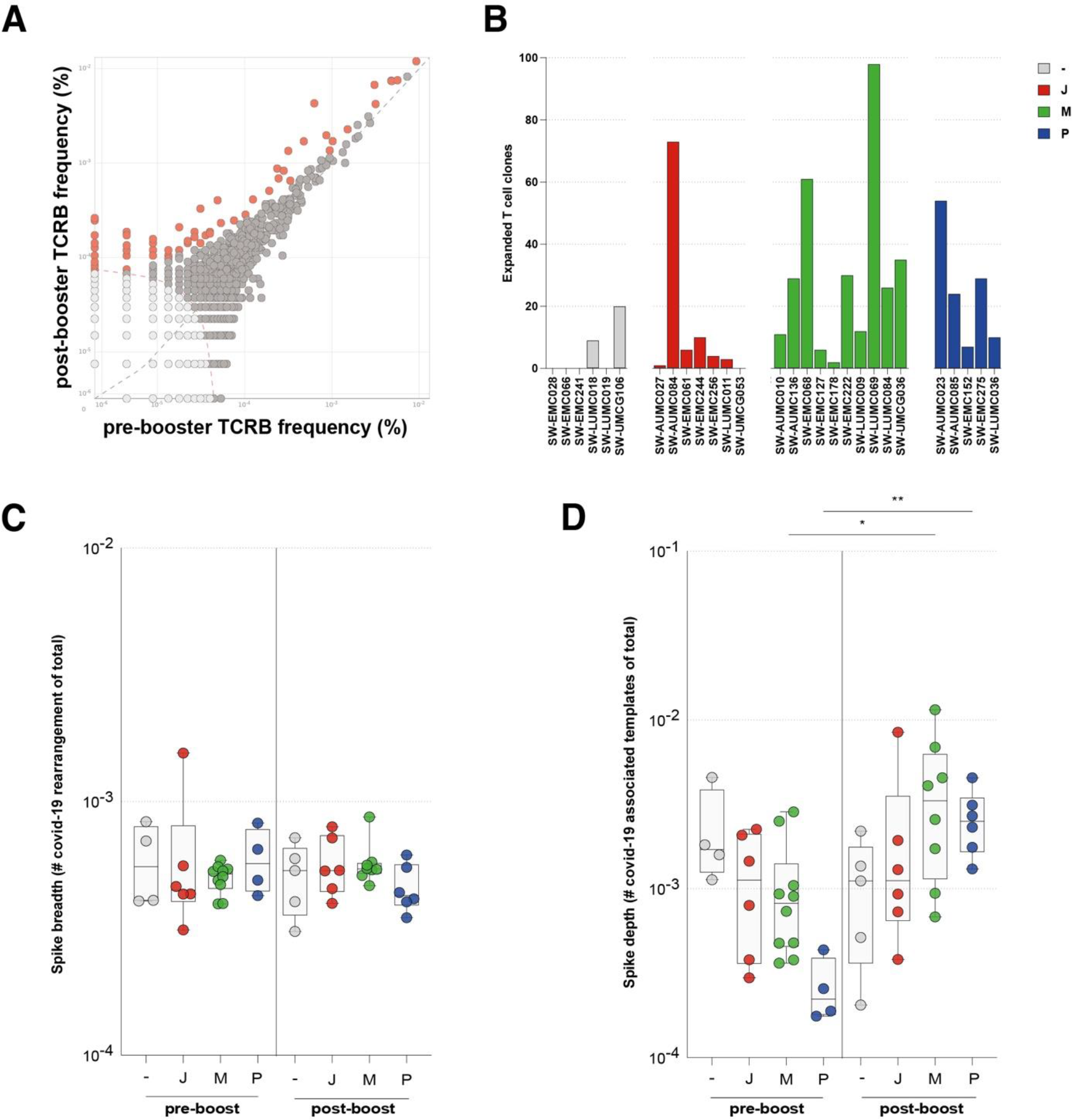
Expansion, breadth and depth of SARS-CoV-2-specific T cell response. **(A)** Representative analysis of clone expansion based on amino acid sequence of the TCR pre- (T1) and post-booster (T2) vaccination. **(B)** Expanded SARS-CoV-2 S-specific T cell clones following vaccination with no boost (grey), Ad26.COV2.S boost (red), mRNA-1273 boost (green), or BNT162b2 boost (blue). **(C)** Breadth and **(D)** depth of the T cell response pre- and post-booster vaccination. - = no boost, J = Ad26.COV2.S, M = mRNA-1273, P = BNT162b2. Symbols represent individual donors. Box plot depicts the median with range (min to max). p <0.05 (*), p <0.01 (**), p <0.001 (***), and p <0.0001 (****).

To identify SARS-CoV-2-specific T cell clones, the TCR sequences were compared to a dataset (the ImmunoCODE MIRA dataset) enriched in COVID-19 cases versus controls *(43)*. This method ensures that clones are specific to SARS-CoV-2 and reduces noise associated with clones that are very frequent or potentially cross-reactive. Breadth (number of unique SARS-CoV-2-specific TCRs) and depth (frequency of SARS-CoV-2-specific TCRs) were calculated for S- **(Fig. 6B and 6C)** and non-S-specific **(Suppl. Fig. 7C and 7D)** T cells. As expected, a dominant S-specific T-cell response was detected, as SARS-CoV-2-infected donors were excluded from analysis **(Suppl. Fig. 7C)**. Interestingly, booster vaccinations did not lead to a significant increase in the breadth of the S-specific T cell response **(Fig. 6B)**. However, booster vaccination with either mRNA-1273 or BNT162b2 led to a significant increase in the depth/frequency **(Fig. 6C)** of the SARS-CoV-2-specific T-cell response, which was not observed after Ad26.COV2.S booster vaccination.

## Discussion

We performed immunological profiling of SARS-CoV-2-specific immune responses, including reactivity to the Delta and Omicron BA.1 variants, after homologous or heterologous booster vaccination of Ad26.COV2.S-primed individuals. We found that Ad26.COV2.S priming provides a solid immunological base for strong and broad SARS-CoV-2-specific immune responses upon subsequent mRNA-based booster vaccination.

Serum samples were collected between August and September of 2021 when Omicron sub-lineages were not prevalent in the Netherlands. To exclude recent infections, a nucleocapsid (N) ELISA was performed on all samples before participants received their booster vaccination *(10)*.

Here, we compared four different booster regimens in a random selection of individuals from the larger SWITCH study *(10)*. Binding antibodies targeting the ancestral SARS-CoV-2, and the Delta and Omicron variants, increased after booster vaccination and levels were highest in participants that received an mRNA-based booster. This is in line with our previous study, in which we found high S1-binding antibodies after heterologous booster vaccination (10). We found that the proportion of RBD-specific memory B cells in blood did not increase after booster vaccination. This indicates that the original Ad26.COV2.S priming induced a sustained RBD-specific memory B cell response and that booster vaccination led to rapid induction of antibody production by memory B cells rather than expansion of SARS-CoV-2-specific memory B cells. As SARS-CoV-2-specific B cells were only measured in blood, it cannot be excluded that a booster may result in the expansion of the memory B cells in the germinal centers.

Antibodies can have a multitude of effector functions, ranging from direct neutralization of the virion to Fc-mediated triggering of cytotoxicity or phagocytosis targeting infected cells and/or cell-free virions, depending on the antibody isotype, glycosylation pattern, and Fc receptor bound *(44)*. Although the majority of the participants in this study developed neutralizing antibodies against the ancestral SARS-CoV-2 and Delta variant (independent of the vaccination regimen), neutralizing antibodies targeting Omicron BA.1 were detected only after mRNA-based booster vaccination, albeit at considerably lower levels compared to the ancestral SARS-CoV-2 *(8)*. Non-neutralizing antibodies are likely to play a role in preventing disease, by targeting virus-infected cells after entry *(26)*. Whereas antibody-mediated neutralization of SARS-CoV-2 is predominantly conferred by binding of antibodies to the receptor binding domain (RBD) of the S protein, non-neutralizing antibodies are less dependent on recognition of epitopes in the RBD and can target the complete S protein as displayed on SARS-CoV-2-infected cells. We assessed Fc-mediated effector functions of antibodies. It was previously hypothesized that these functions might play a role in contributing to protection against COVID-19 *(45, 46)*, but relatively little is known about the impact of Fc-mediated antibody effector functions *(34)*. Novel antigenically distinct SARS-CoV-2 variants, like the Delta variant and Omicron sub-lineages, are partly capable of evading neutralizing antibodies by accumulating mutations in the RBD *(1, 47-49)*. Functional non-neutralizing antibodies are therefore speculated to be less susceptible to immune escape by emerging variants *(25, 34)*. Here, we show an increase in ADCC- and ADCP-mediating antibodies against the ancestral SARS-CoV-2, Delta, and Omicron BA.1 variants, following both homologous and heterologous booster vaccination. Similar to the neutralizing antibody responses, Fc-mediated antibodies were higher following mRNA-based booster vaccination. Although effector functions mediated by non-neutralizing antibodies were also reduced towards the Omicron BA.1 variant, ADCC-mediating antibodies were still clearly detected after mRNA-based booster vaccination. For ADCP we were not able to measure variant-specific responses due to a lack of the required reagents. However, based on the observed correlation between binding, ADCC-mediating and ADCP-mediating antibodies, we expect similar patterns of cross-reactivity.

Similar to non-neutralizing antibodies, SARS-CoV-2-specific T cell responses as well play an important role in reducing COVID-19 severity following re- or breakthrough infection (50). SARS-CoV-2-specific T cells can clear virus-infected cells, contributing to the reduction of virus replication (11). Virus-specific T cells are thought to be long-lived, as these have been detected up to six months after completion of primary vaccination regimens (8), and up to 17 years after SARS-CoV infection (36). T cells can target epitopes dispersed throughout proteins, including conserved epitopes under functional constraints, and are therefore retain cross-reactivity to SARS-CoV-2 variants (9, 30, 40, 41), including the Omicron sub-lineage (7, 8, 51). Here, we show that T cell responses are boosted in Ad26.COV2.S-primed individuals especially after mRNA-based booster vaccination, as measured by both IFN-γ levels and expansion of S-specific T cell clones. Although based on TCRβ sequencing the breadth of the response did not increase after heterologous booster vaccination, reactivity of both CD4 and CD8 T cells with the Delta and Omicron BA.1 variants was retained. No significant increase in CD8 T cell responses was detected following any of the booster vaccinations. This could partly be explained by the fact that CD8 responses are in general difficult to measure from peripheral blood for various reasons. First, the used peptide pools consist of 15-mer peptides which are sub-optimal inducers of CD8 responses that mainly recognize 9- or 10-mer epitopes. Second, the frequency of SARS-CoV-2-specific CD8 T cells in peripheral blood is low and it is thought that CD8 T cells mainly reside in the tissue where they exert their protective function as tissue-resident effector cells.

Currently, several sub-lineages of the Omicron variant are circulating: BA.1-5. Although the BA.1 lineage quickly became dominant upon introduction, it was rapidly replaced by the BA.2 lineage. Both variants have shown a significant escape from neutralizing antibodies *(17, 20, 53, 54)*. Currently, BA.2.12.1, BA.4 and BA.5 are rapidly establishing dominance in different geographical locations *(21-24)*. In our study, we have focused on cross-reactive immune responses to Omicron BA.1, since at the time of the experiments the newer variants were not yet circulating. Based on cross-reactivity with BA.1 and available literature, we expect that non-neutralizing antibodies and T cell responses have at least equal potential for cross-reactivity with these novel immune-evasive variants, based on the targeting of conserved epitopes. Therefore, we speculate that our findings here indicate that heterologous booster vaccination after Ad26.COV2.S priming also induces solid cross-reactive responses to newly emerging variants.

In conclusion, we showed that Ad26.COV2.S priming provides a solid immunological base for SARS-CoV-2-specific immune responses triggered by mRNA-based booster vaccination. Neutralizing antibodies targeting immune-evasive variants were detectable after a mRNA-based booster, and non-neutralizing antibodies and T cell responses to these variants were retained or even boosted. Although there currently is a high prevalence of breakthrough infections with newly emerging Omicron sub-lineages, probably due to the evasion of neutralizing antibodies, the related disease has been reported to be relatively mild *(55)*. Boosting of non-neutralizing antibodies and memory T cells is expected to play an important role in reducing COVID-19 disease severity and could be crucial for vaccine effectiveness in the future *(50, 56)*.

## Methods

### Study design

The SWITCH trial is a single-(participant)-blinded, multi-center, randomized controlled trial among HCWs without severe comorbidities performed in four academic hospitals in the Netherlands (Amsterdam University Medical Center, Erasmus University Medical Center, Leiden University Medical Center, and University Medical Center Groningen), according to the previously described protocol *(58)*. The trial adheres to the principles of the Declaration of Helsinki and was approved by the Medical Research Ethics Committee from Erasmus Medical Center (MEC 2021-0132) and the local review boards of participating centers. All participants provided written informed consent before enrollment.

### Participants

For analysis of humoral and cellular immune responses, 60 donors were randomly selected, taking into account whether sufficient material was available. Participants randomly selected for immunological profiling received a priming vaccination with Ad26.COV2.S, followed by a booster vaccination with Ad26.COV2.S, mRNA-1273 or BNT162b2 after ±95 days (N=15 per group). This differs from the complete original study group, in which the participants received their second vaccination ±84 days after priming with Ad26.COV2.S. As a control group, Ad26.COV2.S primed individuals that were not boosted were included (N=15). Blood samples were collected at day 0 (pre-booster) and day 28 (post-booster), also for the non-boosted control group **(Suppl. Fig. 1A)**.

### Statistical analysis

The baseline characteristics in each group (Ad26.COV2.S/no boost, Ad26.COV2.S/Ad26.COV2.S, Ad26.COV2.S/mRNA-1273, and Ad26.COV2.S/BNT162b2) are described in **Table 1**. Categorical variables are presented as numbers and percentages (%). Differences between the groups were compared with the use of Fisher’s exact test. Continuous variables were presented as medians and interquartile ranges, and between group differences were compared with the use of the Kruskal-Wallis test. We used Mann-Whitney U tests to assess the differences for the following four comparisons: Ad26.COV2.S boost with no boost, Ad26.COV2.S boost with BNT162b2 boost, Ad26.COV2.S with mRNA-1273 boost, and the BNT162b2 boost with an mRNA-1273 boost. For comparing variant responses (ancestral virus, Delta variant and Omicron BA.1 variants) within groups (no boost, Ad26.COV2.S, BNT162b2, or mRNA-1273 boost) we used a Wilcoxon Signed matched-pairs signed rank test. To examine associations between two continuous variables, we estimated a Pearson’s correlation coefficient. A p-value <0.05 was considered statistically significant.

### PBMC and serum isolation

Blood was collected in vacutainer® SST tubes (BD), serum was obtained and stored at -20°C for further experiments. PBMC were isolated from blood and collected in vacutainer tubes containing lithium heparin as anticoagulant by density gradient centrifugation with Lymphoprep™ (Stemcell Technologies) in 50mL SepMate™ collection tubes (Stemcell Technologies) according to manufacturer’s instructions. Briefly, blood was diluted in phosphate buffered saline (PBS), loaded onto Lymphoprep™ and PBMCs were separated by centrifugation at 2000g for 15 minutes. PBMCs were washed 3 times in PBS, counted and frozen in 90% fetal bovine serum (FBS) with 10% DMSO (Honeywell) in liquid nitrogen.

### Detection of S1-specific binding antibodies

Serum samples were tested for anti-S1 immunoglobulin (Ig)G antibodies using a previously validated Liaison SARS-CoV-2 TrimericS IgG assay (DiaSorin, Italy) *(8, 30)*. The lower limit of detection (LLoD) was set at 4.81 binding arbitrary units (BAU)/mL and the responder cut-off at 33.8 BAU/mL. The assay was performed according to manufacturer’s instructions.

### Virus neutralization assay (PRNT50)

Serum samples were tested for the presence of neutralizing antibodies against ancestral SARS-CoV-2, and the Delta and Omicron (BA.1) variants in a plaque reduction neutralization test (PRNT) as previously described *(8, 30, 59)*. Viruses were cultured from clinical material, sequences were confirmed by next-generation sequencing: D614G (ancestral, GISAID: hCov-19/Netherlands/ZH-EMC-2498), B.1.617.2 (Delta, GISAID: hCoV-19/Netherlands/NB-MVD-CWGS2201159/2022), and B.1.1.529 (Omicron BA.1, GISAID: hCoV-19/Netherlands/LI-SQD-01032/2022). The human airway Calu-3 cell line (ATCC HTB-55) was used to grow virus stocks and for PRNT. Calu-3 cells were cultured in OptiMEM (Gibco) supplemented with Glutamax, penicillin (100 IU/mL), streptomycin (100 IU/mL), and 10% fetal bovine serum (FBS). In short, heat-inactivated sera were diluted two-fold in OptiMEM without FBS starting at a 1:10 dilution or in the case of a S1-specific antibody level >2500 BAU/mL starting at 1:80 in 60μL. 400 PFU of each SARS-CoV-2 variant in 60μL OptiMEM medium was added to diluted sera and incubated at 37°C for 1 hour. Antibody-virus mix was transferred onto Calu-3 cells and incubated at 37°C for 8 hours. Cells were fixed in PFA and stained with polyclonal rabbit anti-SARS-CoV-2 nucleocapsid antibody (Sino Biological) and a secondary peroxidase-labeled goat-anti rabbit IgG antibody (Dako). Signal was developed with precipitate-forming 3,3′,5,5′-tetramethylbenzidine substrate (TrueBlue; Kirkegaard & Perry Laboratories) and the number of plaques per well was counted with an ImmunoSpot Image Analyzer (CTL Europe GmbH). The 50% reduction titer (PRNT50) was estimated by calculating the proportionate distance between two dilutions from which the endpoint titer was calculated. Infection controls (no sera) and positive serum control (Nanogram® 100 mg/mL, Sanquin) were included on each plate. A PRNT50 value one dilution step (PRNT50 = 10) lower than the lowest dilution was attributed to samples with no detectable neutralizing antibodies.

### Enzyme-linked immunosorbent assay (ELISA)

Binding antibodies against ancestral SARS-CoV-2, and the Delta and Omicron (BA.1) variants were determined by an in-house developed ELISA as previously described *(59)*. Briefly, ELISA high-binding EIA/RIA plates (Costar) were coated (20ng/well) with baculovirus-generated trimeric prefusion His-tagged S protein from ancestral SARS-CoV-2 (D614G), and Delta and Omicron BA.1 variants (Sino Biological) at 4°C overnight. Next, plates were blocked with blocker blotto buffer in TBS supplemented with 0.01% Tween-20 at 37°C for 1 hour. Consequently, plates were washed and incubated with a 4-fold dilution series of serum starting at a 1:40 dilution at 37°C for 2 hours. Following serum incubation, plates were washed and horseradish peroxidase (HRP)-labeled rabbit anti-human IgG (1:6,000, Dako) was added. Plates were incubated at 37°C for 1 hour, washed and developed with 3,3′,5,5′-tetramethylbenzidine (KPL). Signal was measured at an optical density of 450nm (OD450) using an ELISA microtiter plate reader (infinite F200, Tecan). OD450 signal was corrected by subtracting background signal in the OD620 channel, and a 50% reduction titer was calculated.

### Antibody dependent cellular cytotoxicity (ADCC)

The presence of antibody dependent cell mediated cytotoxicity (ADCC)-mediating antibodies was determined in a previously established assay that measures NK92.05-CD16 cell degranulation *(60)*. In short, high-binding 96-wells plates (Immunolon) were coated with baculovirus-generated trimeric prefusion His-tagged S protein (200 ng/well) from ancestral SARS-CoV-2 (D614G), and Delta and Omicron BA.1 variants (SinoBiologicals) at 4°C overnight. Plates were blocked, washed and incubated with serum (diluted 1:160 and 1:640) at 37°C for 2 hours. Following serum incubation, plates were washed and 100.000 NK92.05-CD16 cells were added, in combination with CD107a^V450^ (1:100, clone H4A3, BD), Golgistop (0.67μL/mL, BD), and GolgiPlug (1μL/mL, BD). Plates were incubated at 37°C for 5 hours, washed and stained for viable NK cells with CD56^PE^ (1:25, clone B159, BD) and LIVE/DEAD Fixable Aqua Dead Cell (AmCyan, Invitrogen, 1:100). Cells were stained at 4°C for 30 minutes and fixed in Cytofix/Cytoperm (BD Biosciences) at 4°C for 30 minutes. Activated NK92.05-CD16 cells were acquired in a FACSLyric (BD) and identified as CD56^+^CD107a^+^ cells. Gating strategy is depicted in **Suppl. Fig. 8A**. Percentages were corrected by subtracting background measured on PBS-coated plates. Two independent experiments were performed, one at a serum dilution of 1:160 **(Suppl. Fig. 8B)** and another at 1:640 **(Suppl. Fig. 8C)** because some samples showed a prozone effect at a 1:160 dilution **(Suppl. Fig 8D and 8E)**, which gave an underrepresentation of the ADCC signal. Average values were used for the main data **(Fig. 2B and 2C)**.

### Antibody dependent cellular phagocytosis (ADCP)

The presence of antibody dependent cellular phagocytosis (ADCP)-mediating antibodies was determined in an assay that measures phagocytosis of S-coated fluorescent beads *(26)*. The monocytic THP-1 cell line (ATCC, TIB-202™) was used to measure ADCP. Briefly, fluorescent Neutravidin beads (FluoSpheres, Life Technologies) were linked to biotin-labeled monomeric S protein from ancestral (D614G) SARS-CoV-2 (Sino Biologicals) by incubating 100μL beads with 100μg protein at 37°C for 2 hours. Sera was added to the S-coated FluoSphere beads in a 4-fold dilution series ranging from a 1:40 to 1:2,560 dilution, or 1:2,560 to 1:163,000 dilution in the case of a S1-specific antibody level >1000 BAU/mL, and incubated at 37°C for 2 hours. 50,000 THP-1 cells were added per well and incubated at 37°C overnight after which FluoSphere bead phagocytosis was measured as PE-positive THP-1 cells by flow cytometry in a FACSLyric (BD). Representative dilution series is shown in **Suppl. Fig. 4A**. ADCP percentages were corrected for PBS control and endpoint titers were determined at an arbitrary cut-off of 20%.

### Detection of RBD-specific B cells by flow cytometry

RBD-specific B cells were measured using fluorescently labeled SARS-CoV-2 RBD-tetramers (SARS-CoV-2 RBD B cell analysis kit, Miltenyi Biotec). In brief, 8-10 × 10^6^ PBMC were incubated with recombinant SARS-CoV-2 RBD-tetramer^PE^ and RBD-tetramer^PE-Vio770^ to stain for RBD-specific B cells. Subsequently, the cells were stained with fluorescently labeled antibodies detecting CD19^APC-Vio770^ (clone LT19), CD27^Vio Bright FITC^ (clone M-T271), IgG^VioBlue^ (clone IS11-3B2.2.3), IgA^VioGreen^ (clone IS11-8E10) and IgM^APC^ (clone PJ2-22H3). Live-dead staining was performed using 7-AAD. Flowcytometry analysis of the whole sample was performed using the FACS Canto II (BD). The proportion of total RBD-specific B cells, RBD-specific memory B cells and RBD-specific IgG memory B cells were determined using FlowJo 10.8.1 (TreeStar, Ashland, OR, USA). The gating strategy is displayed in **Suppl. Fig. 3A**.

### Detection of S-specific T cells by IFN-γ release assay

The presence of SARS-CoV-2-specific T cells was initially measured by a commercially available IFN-ɣ Release Assay (IGRA, QuantiFERON, Qiagen) in whole blood as previously described *(61, 62)*. Briefly, heparinized whole blood was incubated with three different SARS-CoV-2 antigens for 20-24h using a combination of peptides stimulating both CD4 and CD8 T cells (Ag1, Ag2, Ag3, QuantiFERON, QIAGEN). Mitogen-coated tubes were used as positive control and carrier coated tubes were included as negative control. After incubation, plasma was obtained by centrifugation and IFN-ɣ production in response to the antigens was measured by ELISA. Results were expressed in international units (IU) IFN-ɣ/ml after subtraction of the NIL control values as interpolated from a standard calibration curve. Lower limit of detection in this assay was set at 0.01 IU/ml, responder cut-off was 0.15 IU/ml (per manufacturer’s instructions, also used in previous studies *(10)*.

### Detection of S-specific T cells by IFNγ ELISPOT

SARS-CoV-2-specific T cells were measured using IFNγ ELISpot. In short, 17ultiscreen® HTS IP filter plates (Millipore) activated with 35% ethanol were coated with anti-human IFN-γ antibody (1-D1K, Mabtech; 5 µg/mL) and incubated overnight at 4 °C. Next, plates were blocked with X-VIVO (Lonza) medium + 2% Human AB Serum (HS; Sigma). PBMCs were thawed, resuspended in IMDM (Gibco) + 10% FCS, and washed twice. In X-VIVO + 2%HS, PBMCs were brought to a concentration of 4 × 10^6^ cells/mL and rested for 1 hour at 37 °C. SARS-CoV-2 S1 and S2 peptide pools, (JPT Peptide Technologies) consisting of 15-mer peptides overlapping by 11 amino acids that cover the S protein were used for stimulation at a concentration of 0.5 ug/mL. All stimulation were performed in triplicate. 0.4% DMSO (Sigma) was used as negative control and PHA (Remel Europe Ltd; 4 µg/mL) as a positive control. 2 × 10^5^ PBMC were added per well and cultured for 20-24 hours at 37 °C. The next day, ELISpot plates were washed with PBS + 0.05% Tween-20. Anti-human biotinylated IFN-γ antibody (7-B6-1, Mabtech; 1:1000) in 0.05% Poly-HRP buffer (ThermoFisher) was added for 1.5 hours at RT, followed by the addition of Streptavidin poly-HRP (Sanquin; 1:6000) in 0.05% Poly-HRP buffer for 1 hour at RT (in the dark). Spots were developed using TMB substrate (Mabtech). Spot forming cells (SFC) were quantified with the AID ELISpot/Fluorospot reader and calculated to SFCs/10^6^ PBMC. The average of the DMSO negative control was subtracted per stimulation. To define the total S-specific SFC, the sum of SFC of the separate S1 and S2 peptide pools was used. An antigen-specific response of ≥50 SFC/10^6^ PBMCs was considered positive. Samples were excluded when the positive PHA control was negative.

### Detection of S-specific T cells by activation induced marker (AIM) assay

PBMC were thawed in RPMI1640 medium supplemented with 10% FBS, 100 IU/mL penicillin, and 100 IU/mL streptomycin (R10F) and incubated with Benzonase® (50 IU/mL; Merck) at 37°C for 30 minutes. Subsequently, 1×10^6^ PBMC were incubated with in-house developed SARS-CoV-2 peptide pools (15mers with 10 overlap, 1μg/mL per peptide) covering the ancestral, Delta or Omicron BA.1 S protein at 37°C for 20 hours; PBMC were stimulated with an equimolar amount of DMSO as negative control or a combination of PMA (50 μg/mL) and Ionomycin (500 μg/mL) as positive control. Following stimulation, PBMC were stained for surface markers at 4°C for 15 minutes with the following antibodies in their respective dilutions: anti-CD3^PerCP^ (Clone SK7, BD, 1:25), anti-CD4^V450^ (Clone L200, BD, 1:50), anti-CD8^FITC^ (Clone DK25, Dako, 1:25), anti-CD45RA^PE-Cy7^ (Clone L48, BD, 1:50), anti-CCR7^BV711^, anti-CD69^APC-H7^ (Clone FN50, BD, 1:50), anti-CD137^PE^ (Clone 4B4-1, Miltenyi, 1:50), and anti-OX40^BV605^ (Clone L106, BD, 1:25). LIVE/DEAD™ Fixable Aqua Dead Cell staining was included (AmCyan, Invitrogen, 1:100). T cells were gated as LIVE CD3^+^ cells and subdivided into CD4^+^ or CD8^+^ subsets. Memory subsets were identified as either CD45RA^+^CCR7^+^ (naïve, TN), CD45RA^-^CCR7^+^ (central memory, TCM), CD45RA^-^CCR7^-^(effector memory, TEM), or CD45RA^+^CCR7^-^ (terminally differentiated effectors, TEMRA). SARS-CoV-2-reactive T cells were identified as activated T cells after exclusion of TN cells (CD137^+^Ox40^+^ for CD4^+^, or CD137^+^CD69^+^ for CD8+). The gating of subsets and activated cells was set based on the DMSO stimulated sample on a per donor basis. On average, 300,000 cells were acquired on a FACSLyric (BD). Samples with <50,000 counts in the CD3 gate were excluded from analysis. Gating strategy is depicted in **Suppl. Fig. 6A**.

### T-cell receptor variable beta chain sequencing

Genomic DNA (gDNA) was extracted from 1 × 10^6^ PBMC using the DNeasy Blood Extraction Kit (QIAGEN). Depending on the yield, between 12 and 375 μg gDNA was used for immunosequencing of the CDR3 regions of the TCR² chain by the immunoSEQ® Assay (Adaptive Biotechnologies, Seattle, WA). Extracted gDNA was amplified in a bias-controlled multiplex PCR, followed by high-throughput sequencing. Obtained sequences were collapsed and filtered to identify the absolute abundance of each unique TCR TCR² CDR3 region for further analysis. TCR sequences from repertoires were mapped against a set of TCR sequences that are known to react with SARS-CoV-2 by matching on V gene, amino acid sequence and J gene. In brief, these sequences were first identified by Multiplex Identification of T-cell Receptor Antigen Specificity (MIRA, Klinger et al., 2015). The COVID-19 search tool from immunoSEQ was used to identify these SARS-CoV-2-specific T cell clones, and the number of agnostic expanded T-cell clones following booster vaccination was estimated using the differential abundance tool from ImmunoSEQ as previously described *(16)*. Individual responses were quantified by the number and frequency of SARS-CoV-2 TCRs. These were further analyzed at the level of ORF or position within ORF based on the MIRA antigens. The breadth was calculated as the number of unique annotated rearrangements out of the total number of productive rearrangements, while the depth was calculated as the sum frequency of those rearrangements in the repertoire. Two samples were excluded from further analysis due to quality of the sample or gDNA cross-contamination.

## Data Availability

All data needed to evaluate the conclusions in the manuscript is present in the main or the supplementary materials. This work is licensed under a Creative Commons Attribution 4.0 International (CC BY 4.0) license, which permits unrestricted use, distribution, and reproduction in any medium, provided the original work is properly cited. To view a copy of this license, visit https://creativecommons.org/licenses/by/4.0/. This license does not apply to figures/photos/artwork or other content included in the article that is credited to a third party; obtain authorization from the rights holder before using such material.

## Acknowledgements

We acknowledge QIAGEN for supporting the study by providing QuantiFERON SARS-CoV-2 RUO Starter and Extended Packs. QIAGEN had no role in study design, data acquisition and analysis. Assay methodology images were created with Biorender. The NK92.05-CD16 cell line was a kind gift from Kerry S. Campbell at the Fox Chase Cancer Center in Pennsylvania.

## Author contributions

Conceptualization: P.H.M.v.d.K., C.G.v.K., R.D.d.V. Formal analysis: D.G., R.D.d.V., R.S.G.S. Funding acquisition: R.D.d.V., C.G.v.K., P.H.M.v.d.K., R.S.G.S, W.J.R.R., V.A.S.H.D., D.v.B., N.A.K., A.G., D.F.P., L.G.V., A.L.W.H., M.P.G.K., A.S. Investigation: D.G., R.S.G.S, D.v.B., N.A.K., W.J.R.R., K.S.S., S.B., L.G., N.J.N., L.L.A.v.D., M.L. V.A.S.H.D., A.G., D.F.P., L.G.V., A.L.W.H., A.S., A.G., R.L.d.S., M.P.G.K., P.H.M.v.d.K., C.G.v.K., R.D.d.V. Supervision: P.H.M.v.d.K., C.G.v.K., R.D.d.V. Visualization: D.G., R.D.d.V. Writing-original draft: D.G., R.S.G.S., R.D.d.V. Writing: review and editing: all authors reviewed and edited the final version.

## Supplemental Appendix

### SWITCH research group

**Erasmus MC**

N. Tjon

K. van Grafhorst

L.P.M. van Leeuwen

F. de Wilt

S. Scherbeijn

A.C.P. Lamoré

**Amsterdam UMC**

H.M. Garcia Garrido

A.M. Harskamp

I. Maurer

A.F. Girigorie

B. D. Boeser-Nunnink

M.M. Mangas Ruiz

K. A. van Dort

**UMCG**

J.J. de Vries-Idema

J. Zuidema

**LUMC**

J.A. Vlot

P.H. Verbeek –Menken

A.Van Wengen-Stevenhagen

**Supplemental Figure 1:**
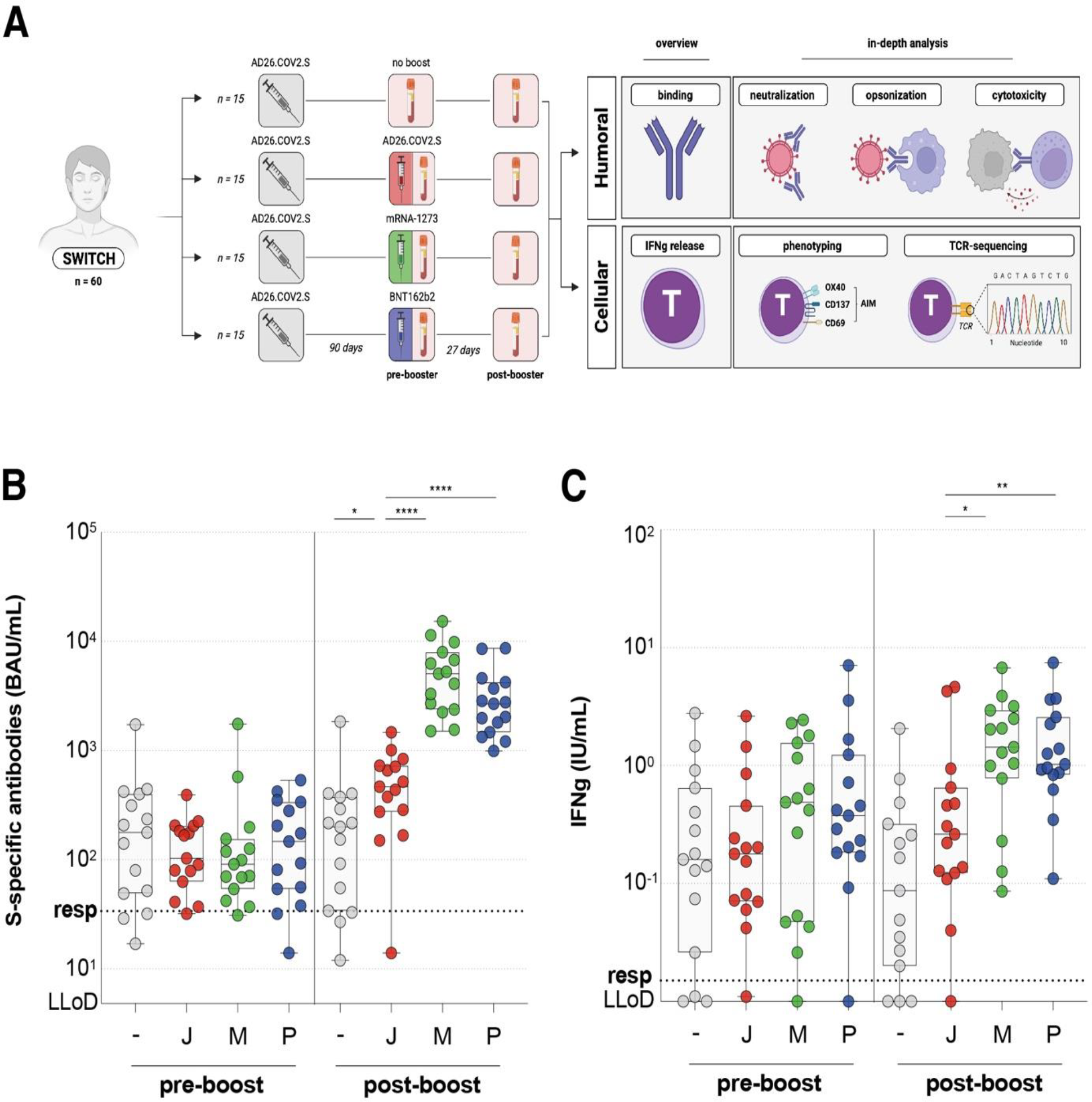
Study design and previously reported binding antibody levels and whole blood T-cell responses. **(A)** Study design of the SWITCH immune profiling analysis. Immune responses in n=60 individuals that received either no (grey), Ad26.COV2.S (red), mRNA-1273 (green), or BNT162b2 (blue) booster vaccination were assessed pre- and post-booster vaccination. **(B)** S-specific antibody levels (BAU/mL) as measured by Liaison and **(C)** IFN-γ levels (IU/mL) as measured by IGRA (Ag2), pre- and post-booster vaccination. For binding antibodies the responder cut-off was set at 33.8 BAU/mL (dotted line), for IGRA at 0.15 IU/mL (dotted line). The lower limit of detection (LLoD) was set at 4.81 BAU/mL for Liaison and 0.01. Symbols represent individual donors. Box plot depicts the median with range (min to max). p <0.05 (*), p <0.01 (**), p <0.001 (***), and p <0.0001 (****).

**Supplemental Figure 2:**
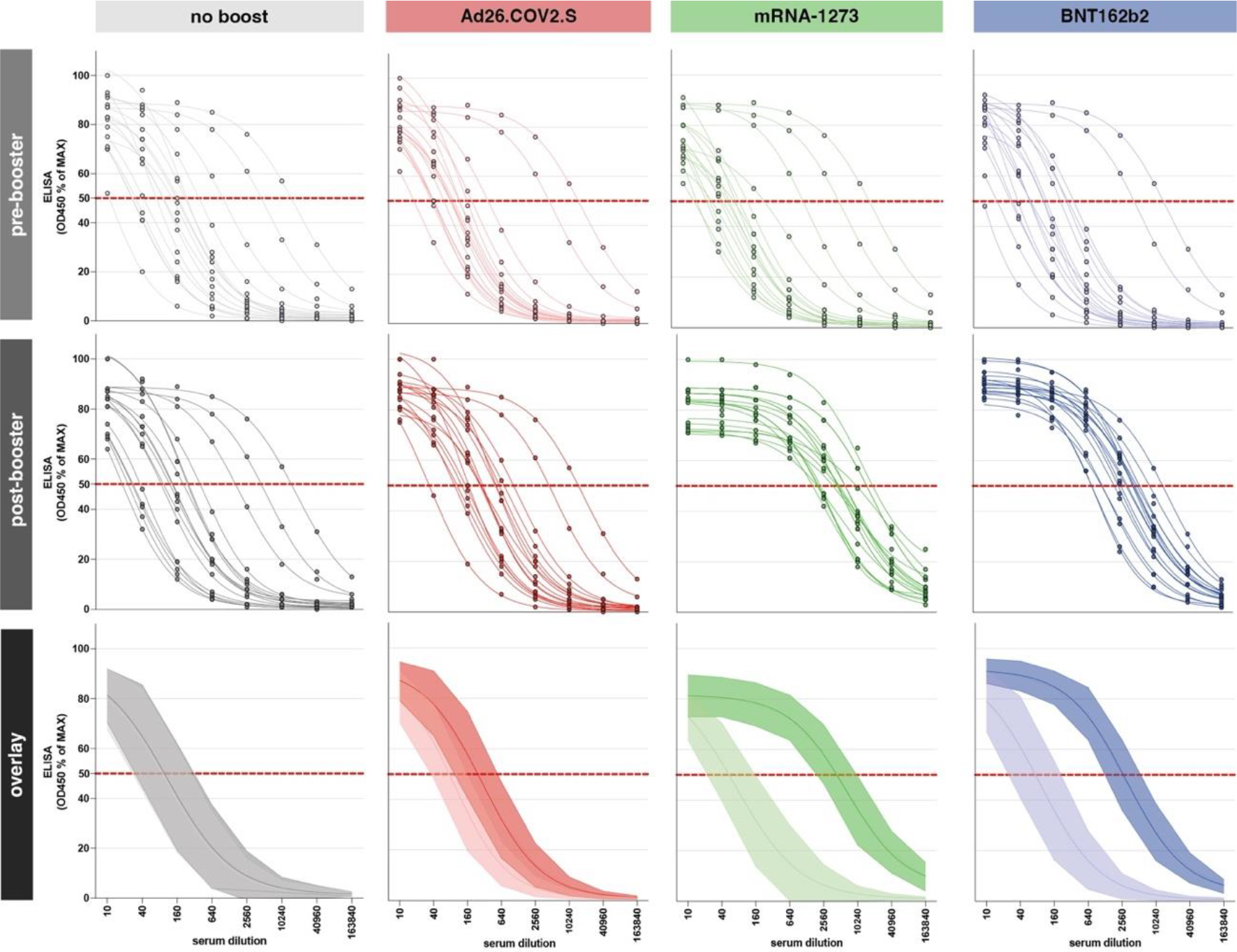
Individual ELISA S-curves per vaccination regimen. log(inhibitor) versus response curves with four parameter variable slopes based on OD450 values % of max per vaccine group, pre-booster vaccination (top), post-booster vaccination (middle), and overlayed depicting the group mean with error bands representing the standard deviation (bottom). Red dotted line indicates the 50% point.

**Supplemental Figure 3:**
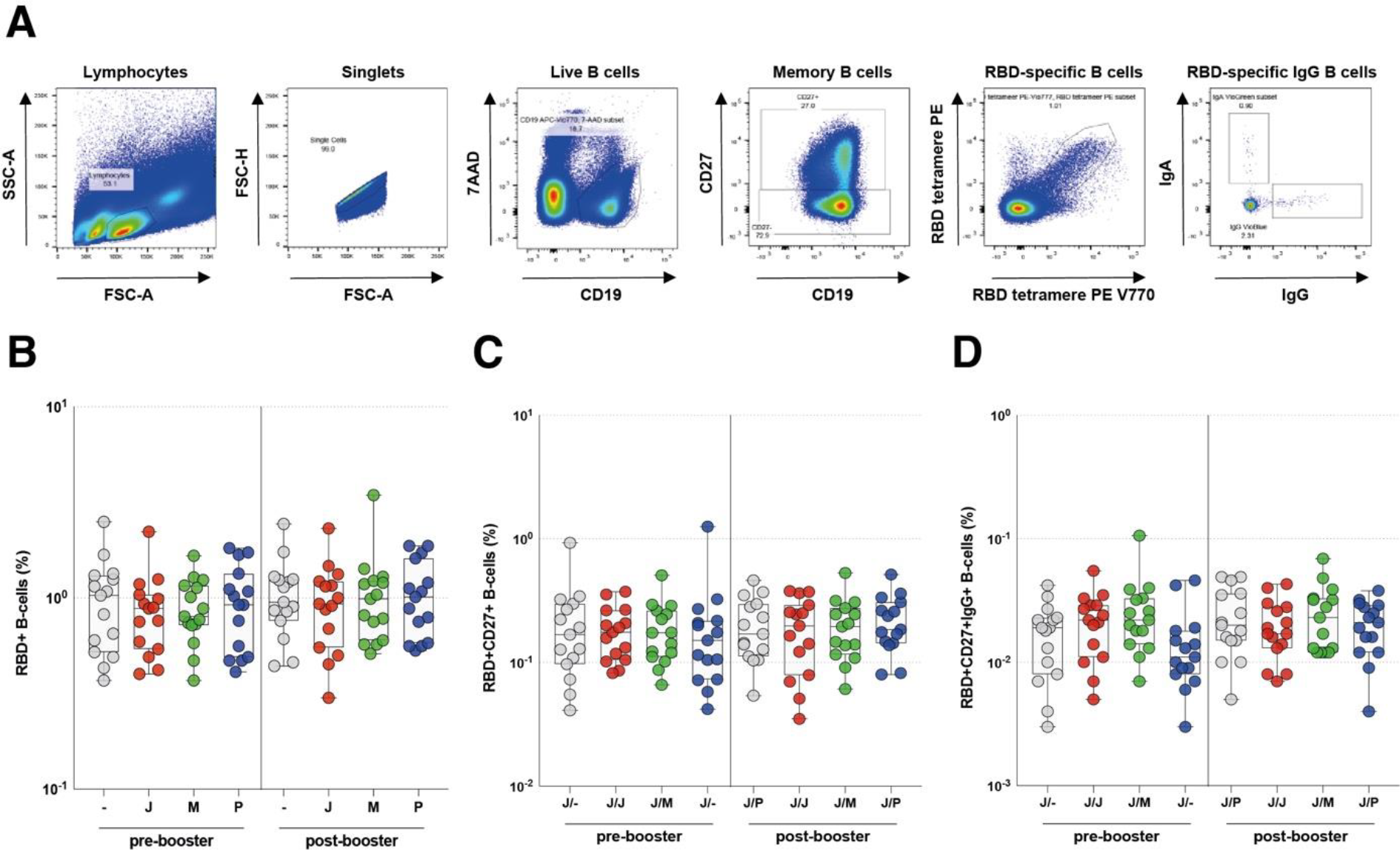
Gating strategy and detection of RBD-specific B-cells. **(A)** Lymphocytes were gated based on the SSC-A and FSC-A and single cells were selected based on FSC-H and FSC-A. Next, LIVE CD19^+^ cells were selected for the analysis of RBD-specific B cells. In addition, the CD27^+^ subpopulation of the CD19^+^ B-cells (memory B cells) and the CD27+ IgG+ subpopulation was selected. In each subpopulation, the proportion of RBD-specific B cells was determined by gating on RBD-tetramer-PE and RBD-tetramer-PE-Vio770 double positive cells. **(B)** Percentage of total RBD-specific B cells, **(C)** RBD-specific memory B cells, and **(D)** RBD-specific IgG memory B cells of total B cells in whole blood after no-boost (grey), Ad26.COV2.S (red), mRNA-1273 (green), or BNT162b2 (blue) booster vaccination pre- and post-booster vaccination.

**Supplemental figure 4:**
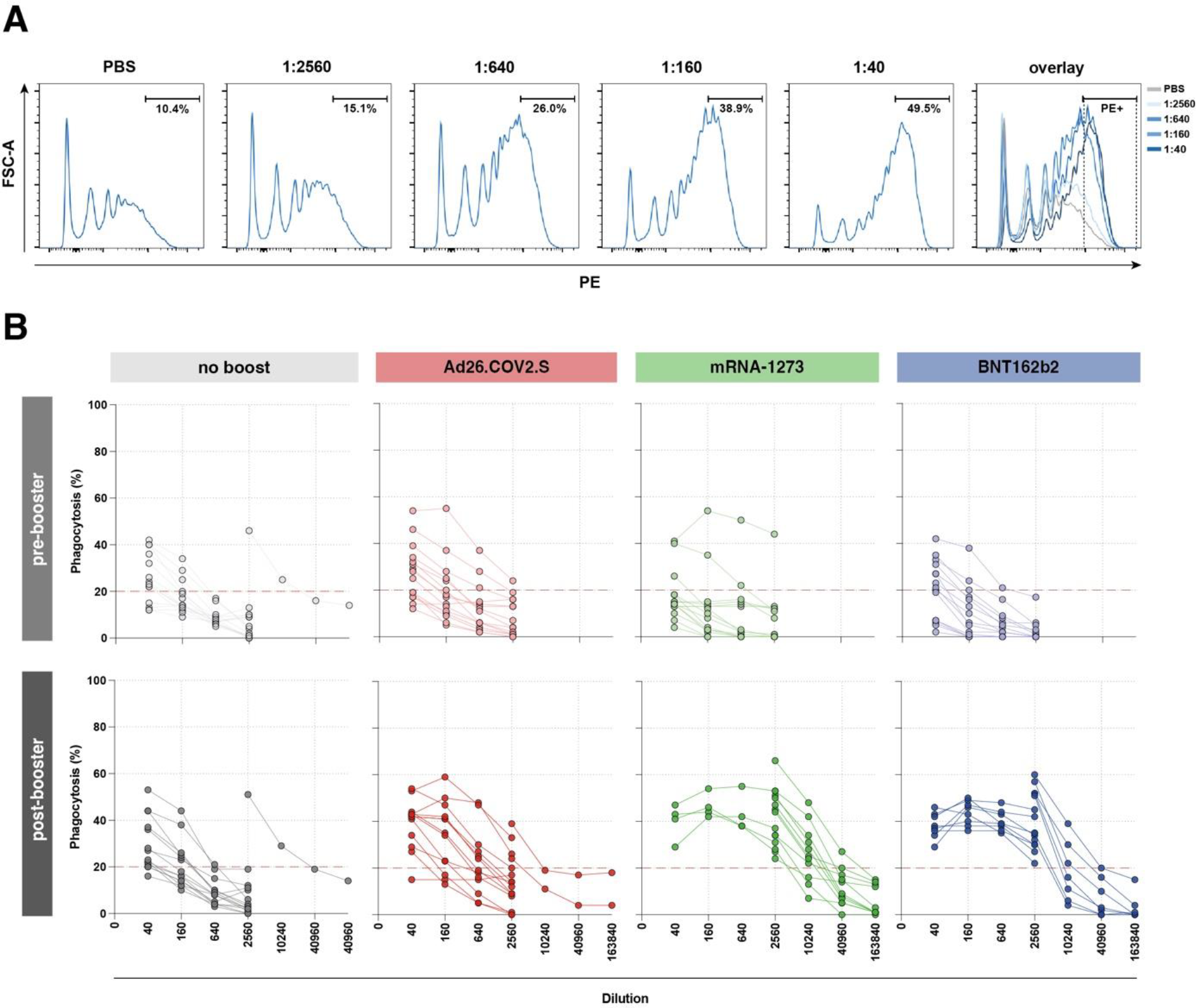
ADCP selection of PE+ cells in a dilution series from one representative sample and individual ADCP S-curves per vaccine regimen. **(A)** Selection of phagocytosing cells. PE signal in PBS control was set at 10% background phagocytosis, and positive sera diluted 1:2560, 1:640, 1:160, and 1:40 from left to right. Overlay depicts PBS control in grey and different dilutions of sera in shades of blue. **(B)** Individual dilution series in ADCP pre- and post-booster vaccination per group: ‘no boost’ (grey), Ad26.COV2.S boost (red), mRNA-1273 boost (green), and BNT162b2 boost (blue). Red dotted line indicates the 20% point dilution used for calculation of ADCP-mediating antibody titers.

**Supplemental figure 5:**
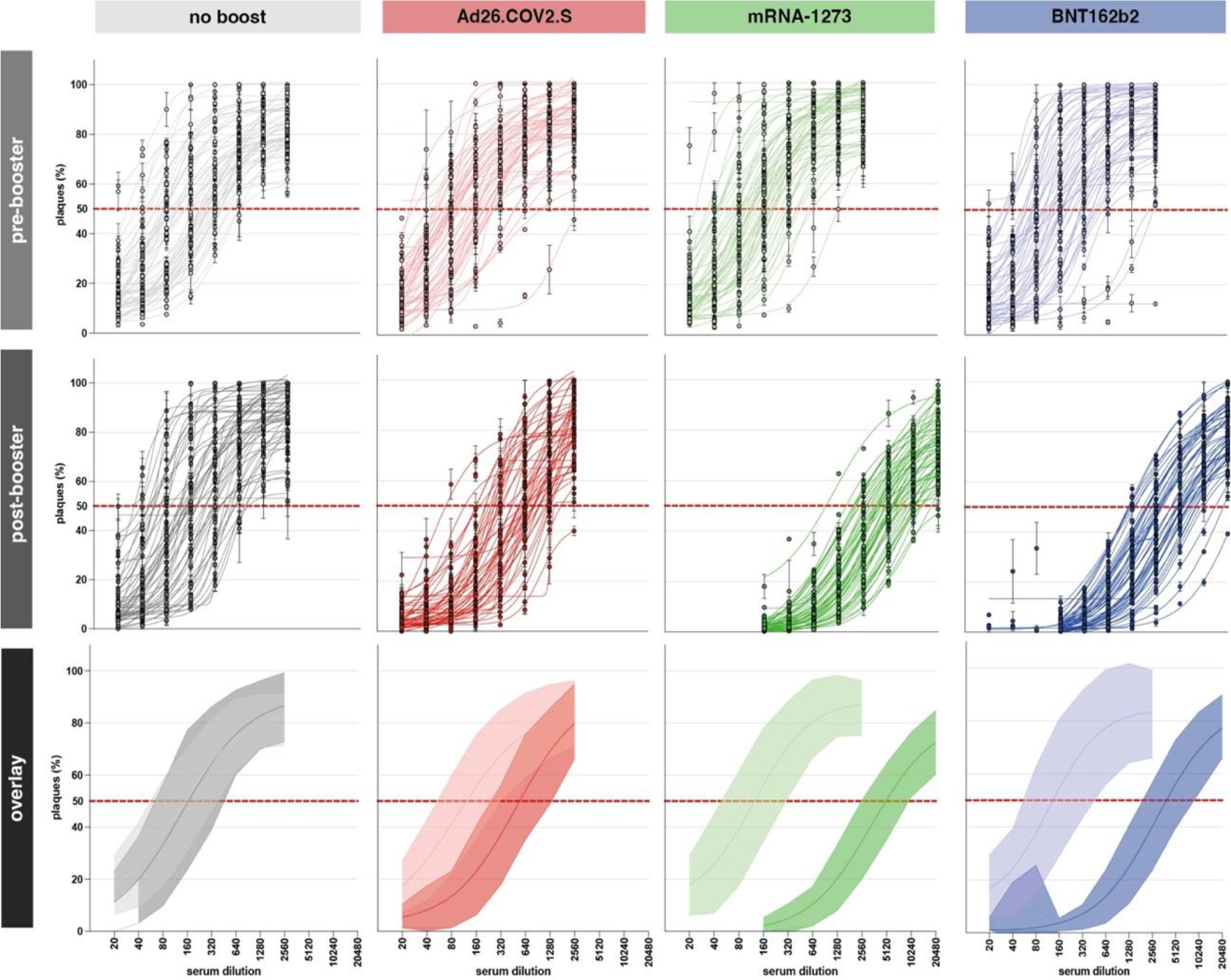
Individual PRNT S-curves per vaccine regimen. log(inhibitor) versus response curves with four parameter variable slopes based on plaque counts compared to the virus control per vaccine group, pre-booster vaccination (top), post-booster vaccination (middle), and overlayed depicting the group mean with error bands representing the standard deviation (bottom). Red dotted line indicates the 50% point.

**Supplemental figure 6:**
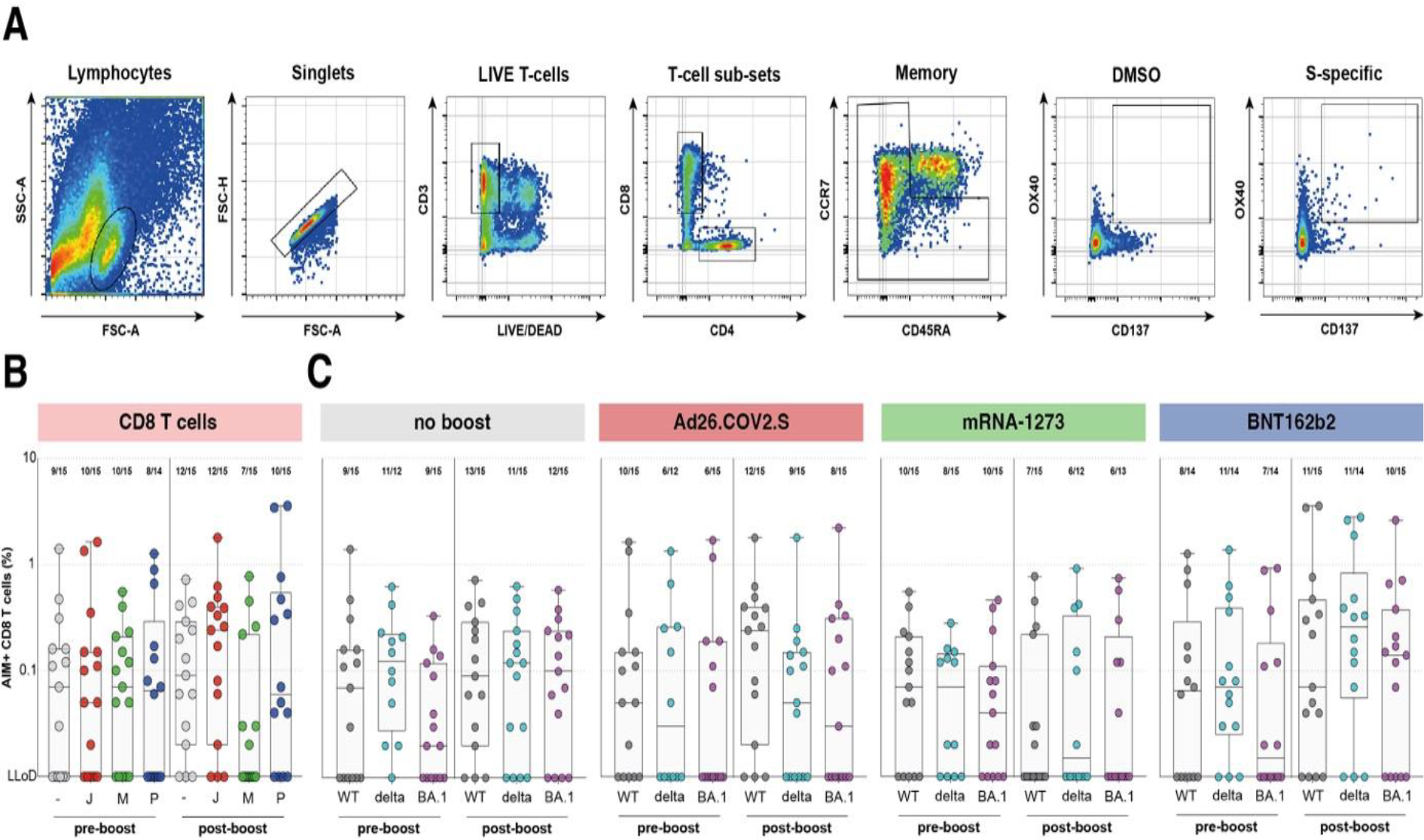
AIM gating strategy and CD8 T cell responses. **(A)** Lymphocytes were gated based on the SSC-A and FSC-A and single cells were selected based on FSC-H and FSC-A. Next, LIVE CD3^+^ cells were selected and sub-divided into CD4^+^ or CD8^+^ T-cells. From each sub-population naïve T-cells are excluded from further analysis based on their expression of CCR7 and CD45RA. Within the memory T-cell population OX40^+^CD137^+^ or CD69^+^CD137^+^ CD4 or CD8 T-cells, respectively, are defined as activated. A representative DMSO stimulated and S stimulated sample is shown. **(B)** AIM expression within the CD8 T cell sub-population against ancestral SARS-CoV-2 and **(C)** variants (Delta and Omicron) per vaccination regimen pre- and post-booster vaccination. Fraction of positive donors is depicted above each group. S = spike protein, - = no boost, J = Ad26.COV2.S, M = mRNA-1273, P = BNT162b2, WT = ancestral virus, del = Delta variant, BA.1 = Omicron BA.1 variant. Symbols represent individual donors. Symbols represent individual donors. Box plot depicts the median with range (min to max).

**Supplemental figure 7:**
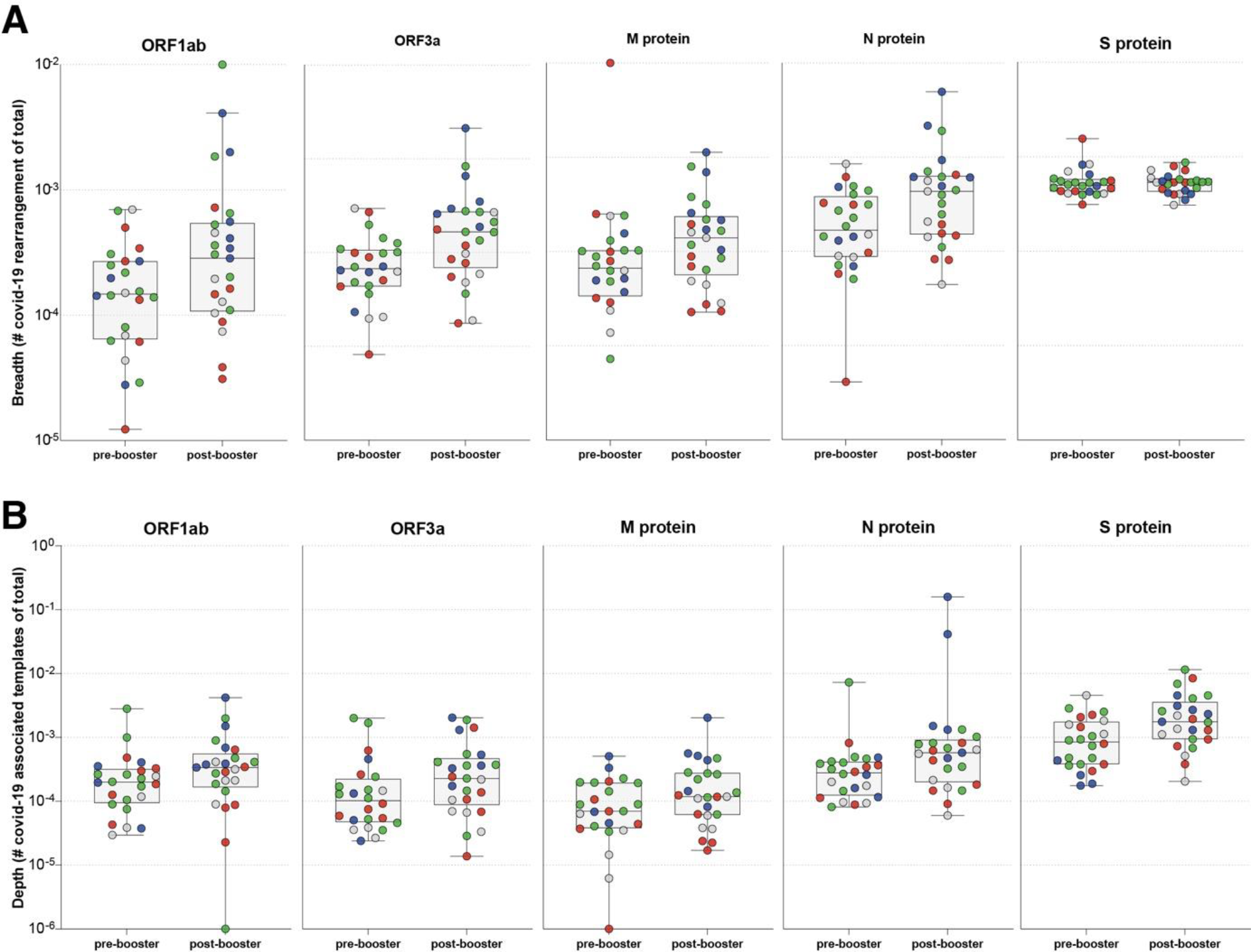
Breadth and depth of SARS-CoV-2-specific T-cell response. **(A)** Breadth and **(B)** depth of the T-cell response to ORF1ab, ORF3a, M protein, N protein, and S protein pre- and post-booster vaccination. Colors represent the vaccine group. Symbols represent individual donors. Box plot depicts the median with range (min to max).

**Supplemental figure 8:**
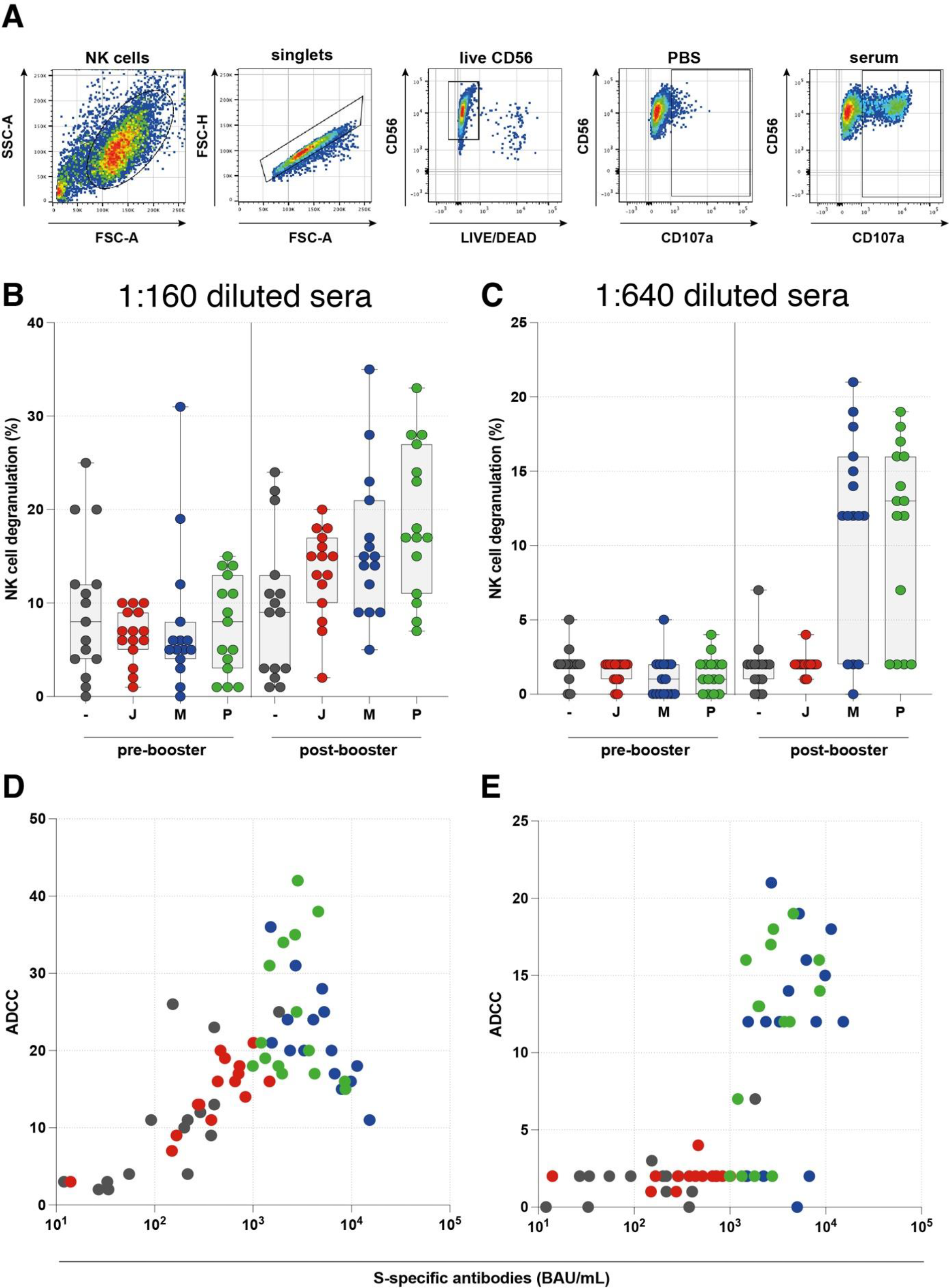
ADCC gating strategy and measurements from two independent experiments at two different serum dilutions. **(A)** Gating strategy for ADCC assay, NK cells were gated on FSC-A and SSC-A, next singlets were selected by FSC-H and FSC-A and LIVE CD56^+^ cells were gated to assess CD107a expression. Representative figure for a PBS and positive serum sample after booster vaccination with mRNA-1273 are shown. **(B)** ADCC data with serum samples diluted 1:160 and **(C)** 1:640. NK cell degranulation is depicted as a % after PBS subtraction pre- and post-booster vaccination after ‘no boost’ (grey), Ad26.COV2.S boost (red), mRNA-1273 boost (green), or BNT162b2 boost (blue). **(D)** correlation of ADCC inducing antibodies and S1-specific binding antibodies at 1:160 dilution and **(E)** 1:640 dilution. Symbols represent individual donors. Box plot depicts the median with range (min to max). p <0.05 (*), p <0.01 (**), p <0.001 (***), and p <0.0001 (****).

